# An international assessment of the COVID-19 pandemic using ensemble data assimilation

**DOI:** 10.1101/2020.06.11.20128777

**Authors:** Geir Evensen, Javier Amezcua, Marc Bocquet, Alberto Carrassi, Alban Farchi, Alison Fowler, Peter L. Houtekamer, Christopher K. Jones, Rafael J. de Moraes, Manuel Pulido, Christian Sampson, Femke C. Vossepoel

## Abstract

This work shows how one can use iterative ensemble smoothers to effectively estimate parameters of an SEIR model with age-classes and compartments of sick, hospitalized, and dead. The data conditioned on are the daily numbers of accumulated deaths and the number of hospitalized. Also, it is possible to condition on the number of cases obtained from testing. We start from a wide prior distribution for the model parameters; then, the ensemble conditioning leads to a posterior ensemble of estimated parameters leading to model predictions in close agreement with the observations. The updated ensemble of model simulations have predictive capabilities and include uncertainty estimates. In particular, we estimate the effective reproductive number as a function of time, and we can assess the impact of different intervention measures. By starting from the updated set of model parameters, we can make accurate short-term predictions of the epidemic development given knowledge of the future effective reproductive number. Also, the model system allows for the computation of long-term scenarios of the epidemic under different assumptions. We have applied the model system on data sets from several countries with vastly different developments of the epidemic, and we can accurately model the development of the COVID-19 outbreak in these countries. We realize that more complex models, e.g., with regional compartments, may be desirable, and we suggest that the approach used here should be applicable also for these models.

## 1. Introduction

We have developed a methodology and software to study the evolution of the COVID-19 pandemic in a country or region and used it to perform an international comparative study across six countries, Argentina, Brazil, England, France, Norway, The Netherlands, four states (New York, California, Alabama, and North Carolina) of the USA, and the province Québec of Canada.

Their diverse geographical locations, including seasonal phase opposition, demography, population densities, and social habits, led the epidemic to evolve in different ways and to impact differently on their very distinct health care systems. Moreover, the implemented counter-measures have been mixed in rigor, timing concerning the epidemic status, and ultimately in their effectiveness. Matters are further complicated by the very diverse data collection protocols, data quality, and degree of accessibility. Accomplishing a comprehensive analysis of such a complicated situation to infer particular aspects of the COVID-19 pandemic and predict its course, requires a joint modeling and data analysis approach and a unified protocol of the study. The scope of this work regards the development of such a common approach. It uses a state-of-art nonlinear ensemble data assimilation method (an iterative ensemble smoother), typically used in geosciences or in petroleum reservoir modeling, to study the COVID-19 pandemic.

Our system combines existing data with a modular SEIR (Susceptible, Exposed, Infectious, Recovered) model with age-classes and additional compartments for sick but quarantined, hospitalized, and dead. We have integrated the SEIR model into an ensemble-based data assimilation framework where the setup resembles methods commonly used for parameter estimation in petroleum reservoir models. The system updates the model state and calibrates its parameters to fit a time series of indirect and noisy observations of deaths, hospitalization, and infected. The current setup constitutes an efficient tool for real-time monitoring and prediction of the COVID-19 epidemic. As opposed to off-line parameter estimation, our system “corrects” the model as soon as new data become available. A key feature of data assimilation is that the model will always track the data to an extent proportional to their assumed accuracy, therefore allowing for a straightforward treatment of incomplete and noisy data, such as those for COVID-19. The ensemble data assimilation provides for assessing the impact of the implemented measures on the magnitude of the effective reproductive number *R*(*t*) as a function of time. Moreover, we can infer the number of undetected infectious, as well as running predictions on the number of fatalities, hospitalizations, and infected, under prescribed social distancing scenarios.

We have used the same model system and ensemble data assimilation method (ESMDA), with similar experimental configurations, for all the cases. However, we have performed different experiments to respond to specific country-dependent questions and their various data provision and characteristics, demonstrating the enormous flexibility of our system and its power. We also present several sensitivity experiments to key data assimilation parameters to address the properties of our approach and its robustness. The model system code and input files for the countries under consideration are available from Github: https://github.com/geirev/EnKF_seir.

Some previous works have used data assimilation for epidemiology, in the context of both variational and Kalman filter-like methods, [8, 53, 31]. Nevertheless, data assimilation, let alone advanced ensemble smoothers, has not been systematically used in epidemiology. Since the advent of the current COVID-19 pandemic, further studies have appeared. These include the work by [60], which, like in the present study, uses an ESMDA approach for parameter estimation in an SEIR model. In [20], a sequential data assimilation method, the Ensemble Kalman Filter, was used to update parameters in a stochastic SEIR model. In contrast, [38] used the Ensemble Adjustment Kalman Filter in combination with a network of SEIR models, simulating different connected “cities” but without age stratification. In the context of variational data assimilation, [58] performed parameter estimation and predictions using a SIR model, while [6] proposed a modified SEIR model that distinguishes between symptomatic and asymptomatic, and uses data assimilation to identify appropriate observing strategies.

The outline of the paper is as follows: Section 2 describes the SEIR model used, and in Section 3, we give a brief introduction to the use of ensemble methods for model calibration. Sections 4–11 present individual results from the different countries, while in Section 12 we make an overall assessment of the obtained results across the modeled countries and states.

## 2. SEIR model with age classes

A simple model for epidemic modeling is the SEIR (Susceptible, Exposed, Infectious, and Recovered) model [9]. For a more realistic description of the COVID-19 epidemic, there is a need for an extended, more sophisticated variant of the SEIR model, and a more convenient formulation is the one shown in Figure 1. Here the populations of susceptible, exposed and infectious are divided into age groups **S**_*i*_, **E**_*i*_, and **I**_*i*_, see, e.g., [13].

**Figure 1.**
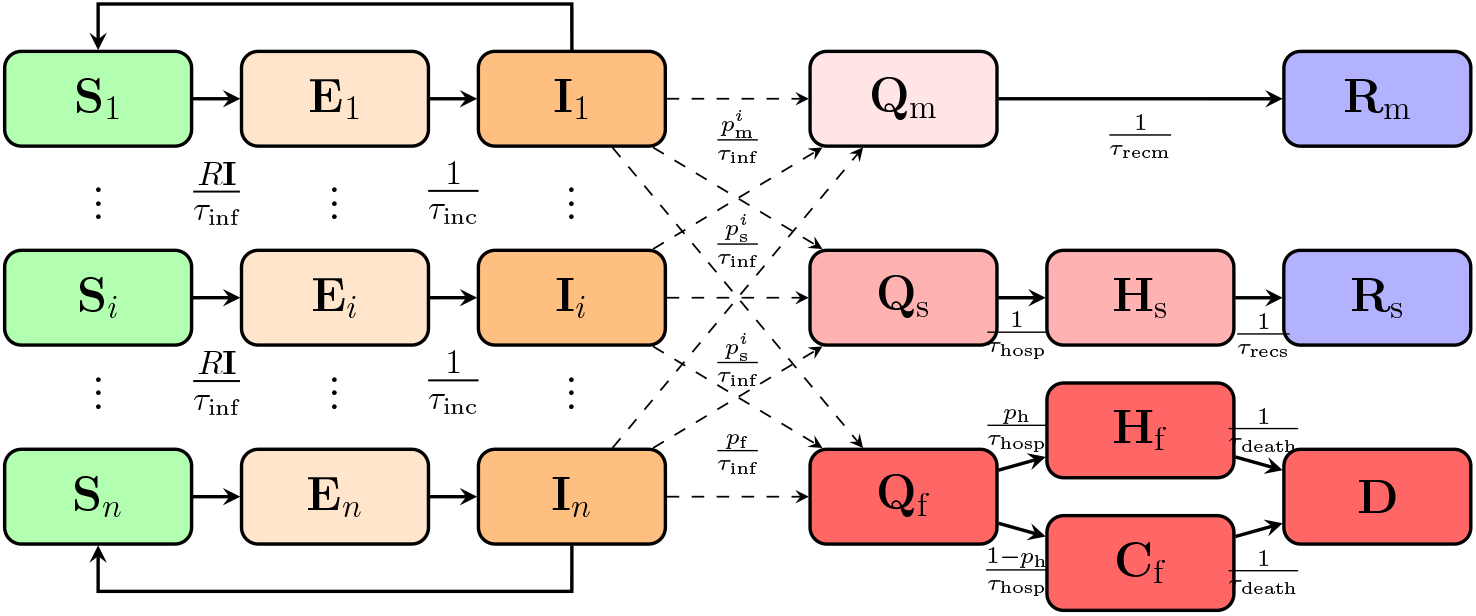
Flow diagram of the SEIR model

The different age groups of infectious **I**_*i*_, transition into the different quarantined groups of sick based on the fractions 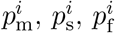, which refer to the portion of patients with mild symptoms, the hospitalized fraction of patients with severe symptoms, and the fraction of fatally ill.

These fractional coefficients sum to one for each age group and allow for specifying how different age groups are affected by the virus. Thus, the model includes different probabilities for dying or being hospitalized dependent on the age group. The specified values for *p*_s_ and *p*_f_ are listed in Table 1.

**Table 1.** The table gives a set of first-guess model parameters.

| Parameter | First guess | Description |
| --- | --- | --- |
| $\tau_{\text{inc}}$ | 5.5 | Incubation period |
| $\tau_{\text{inf}}$ | 3.8 | Infection time |
| $\tau_{\text{recm}}$ | 14.0 | Recovery time mild cases |
| $\tau_{\text{recs}}$ | 5.0 | Recovery time severe cases |
| $\tau_{\text{hosp}}$ | 6.0 | Time until hospitalization |
| $\tau_{\text{death}}$ | 16.0 | Time until death |
| $p_f$ | 0.009 | Case fatality rate |
| $p_s$ | 0.039 | Hospitalization rate (severe cases) |
| $p_h$ | 0.4 | Fraction of fatally ill going to hospital |

Table 2 provides an example set of fractions that illustrates how the older age groups are more severely affected by the COVID-19 virus. The parameter *p*_h_ defines the portions of fatally ill going to the hospital. In Norway, this number is around 0.4, but it varies significantly from country to country.

**Table 2.** The *p* numbers indicate the fraction of sick people in an age group ending up with mild symptoms, severe symptoms (hospitalized), and fatal infection. The population-weighted averages (for the Norwegian population) of the case-fatality rate is 0.0090, and the rate of severe (hospitalized) cases is 0.039.

| Age group | 1 | 2 | 3 | 4 | 5 | 6 | 7 | 8 | 9 | 10 | 11 |
| --- | --- | --- | --- | --- | --- | --- | --- | --- | --- | --- | --- |
| Age range | 0–5 | 6–12 | 13–19 | 20–29 | 30–39 | 40–49 | 50–59 | 60–69 | 70–79 | 80–89 | 90–105 |
| Population | 351159 | 451246 | 446344 | 711752 | 730547 | 723663 | 703830 | 582495 | 435834 | 185480 | 45230 |
| p-mild | 1.0000 | 1.0000 | 0.9998 | 0.9913 | 0.9759 | 0.9686 | 0.9369 | 0.9008 | 0.8465 | 0.8183 | 0.8183 |
| p-severe | 0.0000 | 0.0000 | 0.0002 | 0.0078 | 0.0232 | 0.0295 | 0.0570 | 0.0823 | 0.1160 | 0.1160 | 0.1160 |
| p-fatal | 0.0000 | 0.0000 | 0.0000 | 0.0009 | 0.0009 | 0.0019 | 0.0061 | 0.0169 | 0.0375 | 0.0656 | 0.0656 |

The patients with mild symptoms will recover and transition into **R**_m_ without going to the hospital. The patients with severe symptoms go to the hospital **H**_s_ and, after that, recover into **R**_s_. It is possible to use different definitions of hospitalized, e.g., counting all patients hospitalized because of COVID-19 infections or counting only the ones in ICU by redefining the *p*_s_ parameter.

Finally, the fatally ill patients are either admitted to a hospital **H**_f_ or they are in care homes **C** (or at home) before they end up in the group of dead **D**. This partition of the fatally ill turned out to be very important for most, if not all, of the cases discussed in this paper. The reason is that a significant fraction of the deceased were older people living in care homes. They were usually not admitted to hospitals when they got infected by COVID-19.

### 2.1. Model equations

The model equations are as follows:

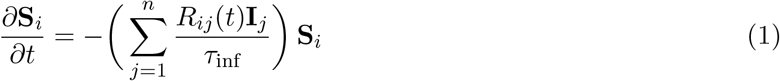

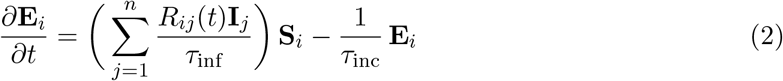

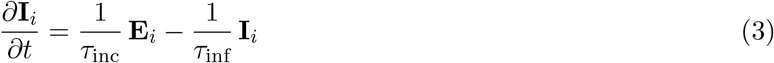

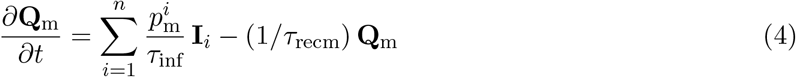

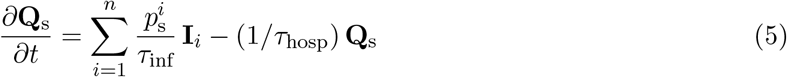

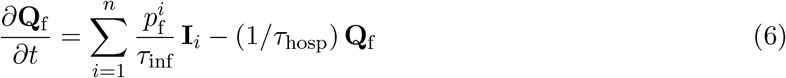

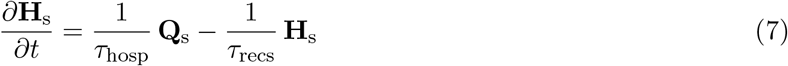

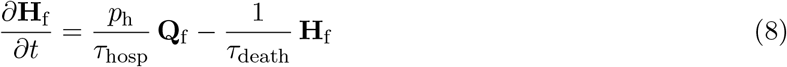

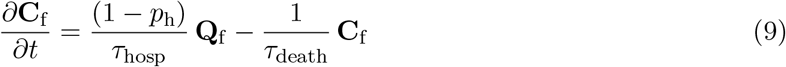

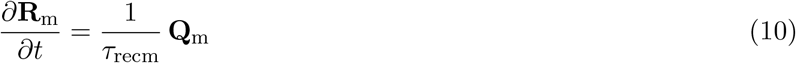

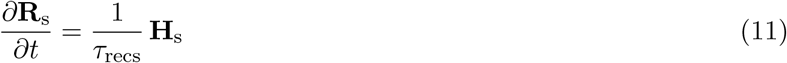

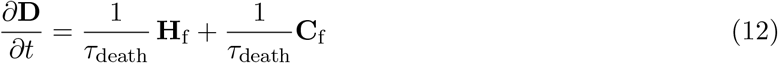

The total population number normalizes these equations. Thus, for this model system, the total population (sum of all model variables, including also **D**) is equal to one and time-invariant. Note also that for the whole population to stay constant in time, the right-hand sides of the equations need to sum to zero.

In the model, there is a flow of people from the **S**_*i*_ groups towards the **R**_m_, **R**_s_, and **D** groups. We prescribe all the transitions in the model, using a rate defined as one over a time scale. There is now a substantial amount of data available, and our data assimilation approach allows us to determine these time scales. Still, we also see that most of them are only marginally updated by assimilating data (i.e., their prior values were fairly correct). Table 1 presents a set of prior values of these time scales.

### 2.2. The effective reproductive number

The most critical parameter in the model is the value of the effective reproductive number *R*(*t*). When *R*(*t*) is higher than one, this leads to an exponential growth of the epidemic, while the epidemic will die out when *R*(*t*) is less than one. As long as *R*(*t*) > 1, the model simulates the development towards the immunity of the population. If at some time, *R*(*t*) becomes less than one and stays less than one for all future times, the epidemic dies out.

When including age groups, it is convenient to use a square matrix **R**(*t*) with a size equal to the number of age groups, which allows for using different reproductive numbers in between the different age groups. It is then possible to include different transmission rates among children (that have fewer symptoms and who may be less infectious) or elders (who got more easily infected in care homes). Also, it allows for simulating increased transmission among children from reopening schools or reduced transmissions among adults who are working from home.

Further, **R**(*t*) can change with time to reflect changes in implemented measures related to social distancing. The equation describing **R**(*t*) in the model is

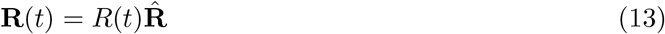

where the scalar function *R*(*t*) is estimated, and 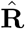 is a prescribed matrix that can differ for up to three different periods. In the model, 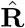 is scaled such that the age-weighted norm 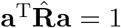. Here the vector **a** contains the fractions of the total population for each age class. Thus, the scalar function *R*(*t*) defines the effective reproductive number, while 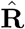 distributes prescribed relative transmissions among the age groups. The default matrix 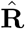 has all elements 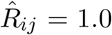, and does not differentiate the transmissions between different age groups.

Initially, *R*(0) equals the basic reproductive number *R*_0_, and the initial transmission rate is *β* = *R*_0_*/τ*_inf_ for an SEIR model without demography [9]. We use effective reproductive numbers *R*(*t*), defined as the average number of secondary cases generated by each infectious person. In the absence of control measures, the effective reproductive number is the fraction of the number of susceptible times *R*_0_. It is possible to reduce the effective reproductive number by introducing intervention measures such as social distancing. In contrast, the basic reproduction number remains unaffected, as it is a measure of the initial rate of infections when there are no interventions [5, page 347].

For the COVID-19 epidemic, it is essential to keep *R*(*t*) below one to avoid exponential growth in the number of infected and thus hospitalized and dead. We can introduce various measures to reduce *R*(*t*) to below one, e.g., immunization by vaccines, reducing social contact, improving hygienic standards by washing hands, and wearing masks.

It is possible to specify different priors for *R*(*t*). We can define *R*(*t*) to be continuously or piecewise continuously varying in time. We can specify an exact prior function, or we can add uncertainty to it, allowing us to update *R*(*t*) using the ensemble methods described below.

### 2.3. Intervention periods

In the model, we have defined three main periods. The first initial period is from the start date of simulation until the introduction of interventions. In this period, we spin up the model from an uncertain initial condition, (size of the population in **I**_*i*_ and **E**_*i*_), and using a significant uncertainty around the prior reproductive number *R*_1_. The second “lockdown” period is from the start of interventions until the “present” time. For this period, we can set a lower prior value of the effective reproductive number, *R*_2_, to reflect the interventions’ expected impact. The final period is for the prediction where we must assume the future distribution for *R*(*t*) around the prior *R*_3_. For the three different periods, we can give various structure matrices 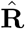.

## 3. Ensemble methods for model calibration

There is a vast literature on the use of ensemble Kalman filter (EnKF) type methods for sequential data assimilation, model parameter estimation, and solving high-dimensional and nonlinear inverse problems. Ensemble data assimilation is now standard and state of the art in a majority of operational prediction systems in the geosciences [14], including weather prediction [32], and petroleum applications [4]. The most popular ensemble-based data assimilation methods build on the Ensemble Kalman Filter (EnKF) originally developed by [27, 28, 21, 22].

Ensemble data assimilation in geosciences and petroleum applications solves the state and parameter estimation problem. The problem, being highly nonlinear, represented a formidable challenge, with a solution leading to a great stream of research and development [7]. We adopt a similar approach for our SEIR model, and thus we benefit from the extensive existing theory and methods initially flourished in geosciences and petroleum research.

Let us write the model Eqs. (1–12) in compact form as

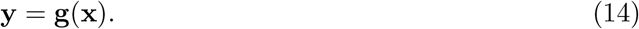

Here, **x** is a vector containing all the uncertain parameters of the model, including initial conditions, and the time-varying *R*(*t*). The predicted measurements **y** relates to the input parameters **x** through the model equations **g**. We have time series of measurements of the number of deaths, the number of people hospitalized, and the number of cases,

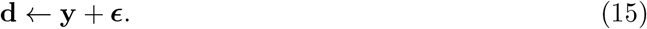

The inverse problem solves for **x** given the predicted measurements, **y**, and the observations **d**. This inverse problem can be framed using Bayes’ theorem [23], as

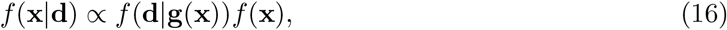

where *f* (**x** |**d**) is the posterior probability density function of the parameters, **x**, conditioned on the data **d**. Equation 16 defines the so-called smoothing problem. Our current approach is to use ensemble methods to approximately solve this equation [23, 7].

Developments originating from the petroleum applications have led to the use of new iterative ensemble smoothers such as the ensemble smoother with multiple data assimilation (ESMDA) by and the ensemble randomized maximum likelihood (EnRML) by [17, 18]. Similar methods have been developed in the geosciences albeit, within a time-sequential context [10, 11]. These methods have become popular for solving inverse problems with moderate nonlinearity. Recently, [51, 26] introduced a new efficient formulation of the EnRML, that searches for the solution in the ensemble subspace and this method is now operational in petroleum applications [26, 25].

Without going into the mathematical details, the ensemble methods work as follows.

1. First sample a large ensemble of realizations of the prior uncertain parameters (e.g., the parameters listed in Table 1, the function *R*(*t*), and the initial infected and exposed **I**_*i*_ and **E**_*i*_), given their prescribed first-guess values and standard deviations.
2. Integrate the ensemble of model realizations forward in time to produce a prior ensemble prediction, which also characterizes the uncertainty.
3. Compute the posterior ensemble of parameters by using the misfit between prediction and observations and the correlations between the input parameters and the predicted measurements.
4. Finally, compute the posterior ensemble prediction by a forward ensemble integration. The posterior ensemble is then the “optimal” model prediction with the ensemble spread representing the uncertainty.

Assuming that an increase in a parameter value yields an increase in a predicted measurement, then there is a positive correlation between the parameter and the prediction. It is then possible to use this correlation to adjust the parameter value such that the posterior model prediction is closer to the observations.

For solving the COVID-19 data assimilation problem, it is convenient to use an ensemble smoother. The main reason is that the time-varying and poorly known reproductive number, *R*(*t*), at a particular time, determines the model predicted deaths and hospitalizations, two to three weeks into the future. A sequential data assimilation method would update the predicted state variables, but not be as suitable for correcting a possible model bias caused by a poor estimate of *R*(*t*) a couple of weeks before.

In this study, we use the ESMDA method because of its simplicity of implementation, and its computational efficiency with large ensemble sizes. For a comprehensive explanation of ESMDA for solving parameter-estimation problems, see [23]. In particular, the ESMDA algorithm solves the Bayesian parameter estimation problem by gradually introducing the information from the measurements to mitigate the impact of nonlinearity. The method handles strong nonlinearity, high-dimensional problems, and a vast number of observations. In the Appendix, we have added a brief mathematical description of the ESMDA algorithm and commented on its convergence related to the number of steps and ensemble size.

## 4. A case study for Norway

In Norway, the Government acted quickly and imposed a lockdown, closing all kindergartens, schools, universities, and all noncritical functions on March 15th. Several companies ordered their staff to work from home. All restaurants and bars closed. Still, society remained partly open. People were allowed to go hiking and training outside as long as there was no physical contact between them. The interventions did not confine Norwegians to their homes as in several other countries, and the psychological impact has probably been less severe. Considerable controversy surrounded the ban against going to cabins in the mountains or along the coast. But this ban most likely reduced the spread of the virus between different regions. Norway has a relatively small population of a little more than five million people spread over a large geographical area, thus keeping a distance is not difficult.

The interventions came after about two to three weeks with sporadic imports of cases, mainly through people returning from skiing vacations in Austria and Italy. In the days before the closedown, there was an increasing number of cases where the infection’s origin was unknown. Thus, it was clear that the virus was spreading through the population. The initial lockdown was active for five weeks, until April 20th, when the kindergartens reopened. The schools opened for the first four cohorts one week later on April 27th. During April, Norway had a steady reduction in the number of hospitalized COVID-19 patients from a maximum of 322 patients on March 30th. The government, therefore, decided to reopen society gradually, starting with kindergartens and schools. During this period, the number of deaths and the number of new cases per day did not increase. In the following weeks, some restaurants started opening again, but with a minimum distance between tables and no accumulation of people. As of June 6th, Norway had experienced only three deaths in the previous two weeks, only 24 COVID-19 patients were in the hospital, and the number of newly detected cases per day was around ten. Norway was able at this point to test anyone with a symptom and run active contact tracing related to all positive cases. Thus, the general opinion was that Norway had control of the epidemic.

The initial model development was motivated by the need for monitoring and predicting the epidemic in Norway. In particular, we believed that the introduction of ensemble-based data-assimilation methods for model calibration would lead to an ideal tool for making short term predictions, evaluating scenarios, and estimating the impact of different interventions, or their releases, on the effective reproductive number. We communicated the model scenarios to Norwegian authorities to provide decision support related to managing the interventions.

The age groups were added to the model to afford a simulation of the impact of the reopening of schools and kindergartens. By increasing the transmissions among children who would be interacting at school, and introducing a slight increase among the children and their parents, it was reasoned that this would lead to a higher effective reproductive number. If the effective reproductive number were to get above one, the spreading would be unstable. Thus, excessive transmissions among children would have to be balanced by less spread among the adult population.

### 4.1. Impact of gradually re-opening schools

On April 14th, we used the ensemble method presented in Sec. 3 with the model described in Sec. 2 to run sensitivity simulations for assessing the impact of gradually reopening children’s schools and kindergartens from April 20th. The age-based model considered 11 age groups as defined in Table 2.

The actual values in **R**(*t*) were initially the same between all age groups until April 20th when schools and kindergartens reopened. Then, we introduced the 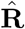 matrix from Table 3, used in the simulation after letting children back to kindergartens and schools. The matrix allowed for using different transmission factors between different age groups. On the diagonal, the value gives the transmission of disease within the same age group. The off-diagonal terms are the transmissions between age groups. We assumed that open kindergartens and schools would lead to significant spreading of the virus within these groups. We also included an increased transmission between parent groups and children. Note that the weight-averaged norm of this matrix is one, so it does not contribute to the effective reproductive number, *R*(*t*).

**Table 3.** Norway: This 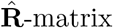 increases transmissions among children after opening kindergartens and schools on April 20th.

| Age groups | 1 | 2 | 3 | 4 | 5 | 6 | 7 | 8 | 9 | 10 | 11 |
| --- | --- | --- | --- | --- | --- | --- | --- | --- | --- | --- | --- |
| 1 | **3.3** | 1.8 | 1.8 | 1.3 | 1.3 | 1.0 | 0.9 | 0.9 | 0.9 | 0.9 | 0.9 |
| 2 | 1.8 | **3.3** | 1.8 | 1.3 | 1.3 | 1.3 | 0.9 | 0.9 | 0.9 | 0.9 | 0.9 |
| 3 | 1.8 | 1.8 | **0.9** | 0.9 | 0.9 | 0.9 | 0.9 | 0.9 | 0.9 | 0.9 | 0.9 |
| 4 | 1.3 | 1.3 | 0.9 | **0.9** | 0.9 | 0.9 | 0.9 | 0.9 | 0.9 | 0.9 | 0.9 |
| 5 | 1.3 | 1.3 | 0.9 | 0.9 | **0.9** | 0.9 | 0.9 | 0.9 | 0.9 | 0.9 | 0.9 |
| 6 | 1.0 | 1.3 | 0.9 | 0.9 | 0.9 | **0.9** | 0.9 | 0.9 | 0.9 | 0.9 | 0.9 |
| 7 | 0.9 | 0.9 | 0.9 | 0.9 | 0.9 | 0.9 | **0.9** | 0.9 | 0.9 | 0.9 | 0.9 |
| 8 | 0.9 | 0.9 | 0.9 | 0.9 | 0.9 | 0.9 | 0.9 | **0.9** | 0.9 | 0.9 | 0.9 |
| 9 | 0.9 | 0.9 | 0.9 | 0.9 | 0.9 | 0.9 | 0.9 | 0.9 | **0.9** | 0.9 | 0.9 |
| 10 | 0.9 | 0.9 | 0.9 | 0.9 | 0.9 | 0.9 | 0.9 | 0.9 | 0.9 | **0.9** | 0.9 |
| 11 | 0.9 | 0.9 | 0.9 | 0.9 | 0.9 | 0.9 | 0.9 | 0.9 | 0.9 | 0.9 | **0.9** |

A base experiment was run fitting model parameters to the total observed deaths and the current number of hospitalized using data until April 14th. We configured the model with a spinup period before the start of the interventions on March 15th. In this spinup period, we used a prior value *R*_1_ = 4.5 and a standard deviation of 0.2. From March 15th to April 14th, during the interventions, we used a prior value of *R*_2_ = 0.80 and a standard deviation of 0.15. The model parameters, including *R*(*t*), were updated using the ESMDA method.

We communicated three prediction scenarios using the 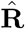 matrix in Table 3 from April 20th, and with different prior values of *R*(*t*) in the prediction phase, *R*_3_ = 0.8, *R*_3_ = 1.0, and *R*_3_ = 1.2, all with a standard deviation of 0.15. Figure 2 presents the results of these cases, with increasing value of *R*_3_ for the prediction, from top to bottom.

**Figure 2.**
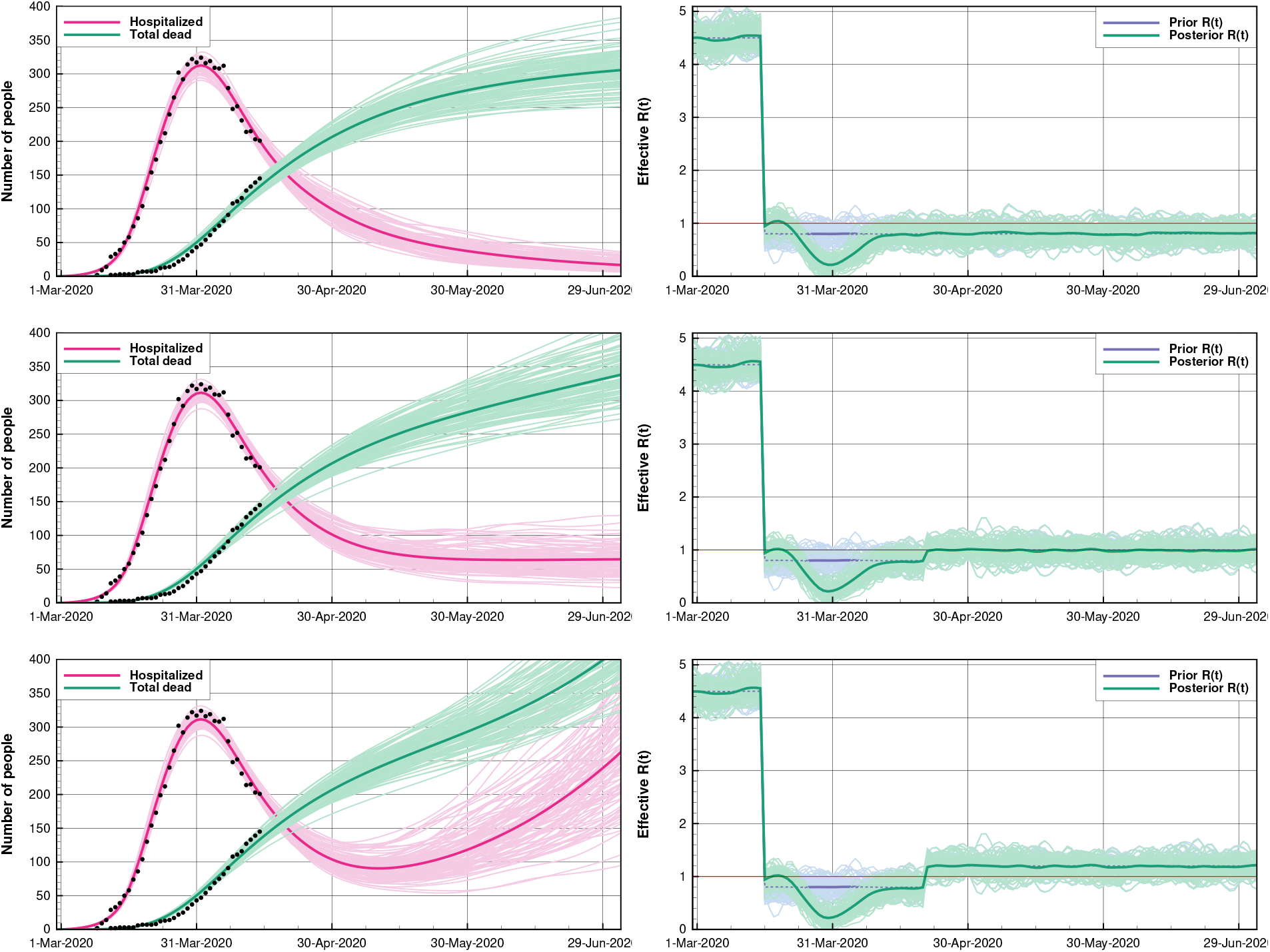
Norway: This figure presents a summary of scenarios related to opening up kindergartens and schools on the 20th of April. The left plots show the ensemble means and the 100 first ensemble realizations, for the number of hospitalized and the accumulated amount of deaths for different scenarios of future *R*(*t*) = 0.8, 1.0, and 1.2. The right plots show the prior and posterior ensembles of *R*(*t*).

It turned out we were too pessimistic in the predictions. The lockdown in Norway was effective and nearly stopped further spreading of the virus during the five weeks between March 15th and April 20th. Thus, we ran scenarios with values of *R*_3_ that were too high, and predicted a more severe development of the epidemic than actually occured.

More important than predicting the exact number of fatalities and hospitalized, was to illustrate the qualitative evolution of the pandemic in the three cases when the effective reproductive number is less than one (stable), equal to one (neutral), or larger than one (unstable). The three scenarios with different values of *R*_3_ result in vastly different future predictions, and it is essential to keep *R*(*t*) less than one.

Small changes in the interventions can lead to an *R*(*t*) that is larger than one, and two-three weeks later, a bloom of new cases will result. Thus, at an early stage of the pandemic, it is essential to halt the spreading of the virus abruptly and obtain control of the active cases. After that, it is possible to open society gradually while monitoring the impact on the reproductive number. Finally, from the 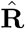 matrix in Table 3, we see that it is possible to balance increased spreading within some age groups by fewer transmissions in other age groups.

A conclusion made based on these simulations was the following: *The opening of children’s schools and kindergartens would likely yield growth in new cases for the young age groups. The virus then spreads to their parents, teachers, and the rest of the community. A continuous lockdown in the remainder of society might have stabilized the growth, but the measures must be sufficient, or we might have experienced exponential growth again. With open schools, it is not likely that the rest of the society would manage to keep R*(*t*) *below one due to the additional commuting and people going back to work. We suggested that the extreme transmission rate for the Coronavirus would make the opening of parts of society very risky. We would likely experience multiple new local exponential blooms of infected people*. We have later experienced a few local “blooms” and an increase in *R*(*t*), but, as will be discussed below, the partial and gradual reopening strategy has worked out very well.

### 4.2. Recent scenarios for Norway

A more recent scenario includes all the data until the end of May, and Figure 3 presents the results. The left plots show an optimized case and an online estimation of *R*(*t*) during the lockdown period. In this case, after March 15th, we used a prior value of *R*(*t*) = 0.8 with a standard deviation of 0.15. The interventions implemented on March 15th led to an immediate reduction of *R*(*t*) in the following week, and *R*(*t*) reached a minimum of 0.3 to 0.4 on March 31st. During April, there was the gradual restoration of *R*(*t*) towards its prior value of 0.8. Using data up to the end of May, we would expect to see an impact on *R*(*t*) through mid-May. Thus, to complement this discussion, we re-ran the experiment but using a prior value of *R*(*t*) = 0.6. The plots in the right column of Figure 3 show the results, and we see that there was indeed a positive uptick of *R*(*t*) until mid-May. It turned out that the prior *R*(*t*) in the first case was too close to the correct value for May. These experiments confirm that new measured deaths and hospitalizations impact the updates of *R*(*t*) about two weeks earlier. Thus, infections must have happened about two weeks earlier since it takes some time before infected people get hospitalized or die.

**Figure 3.**
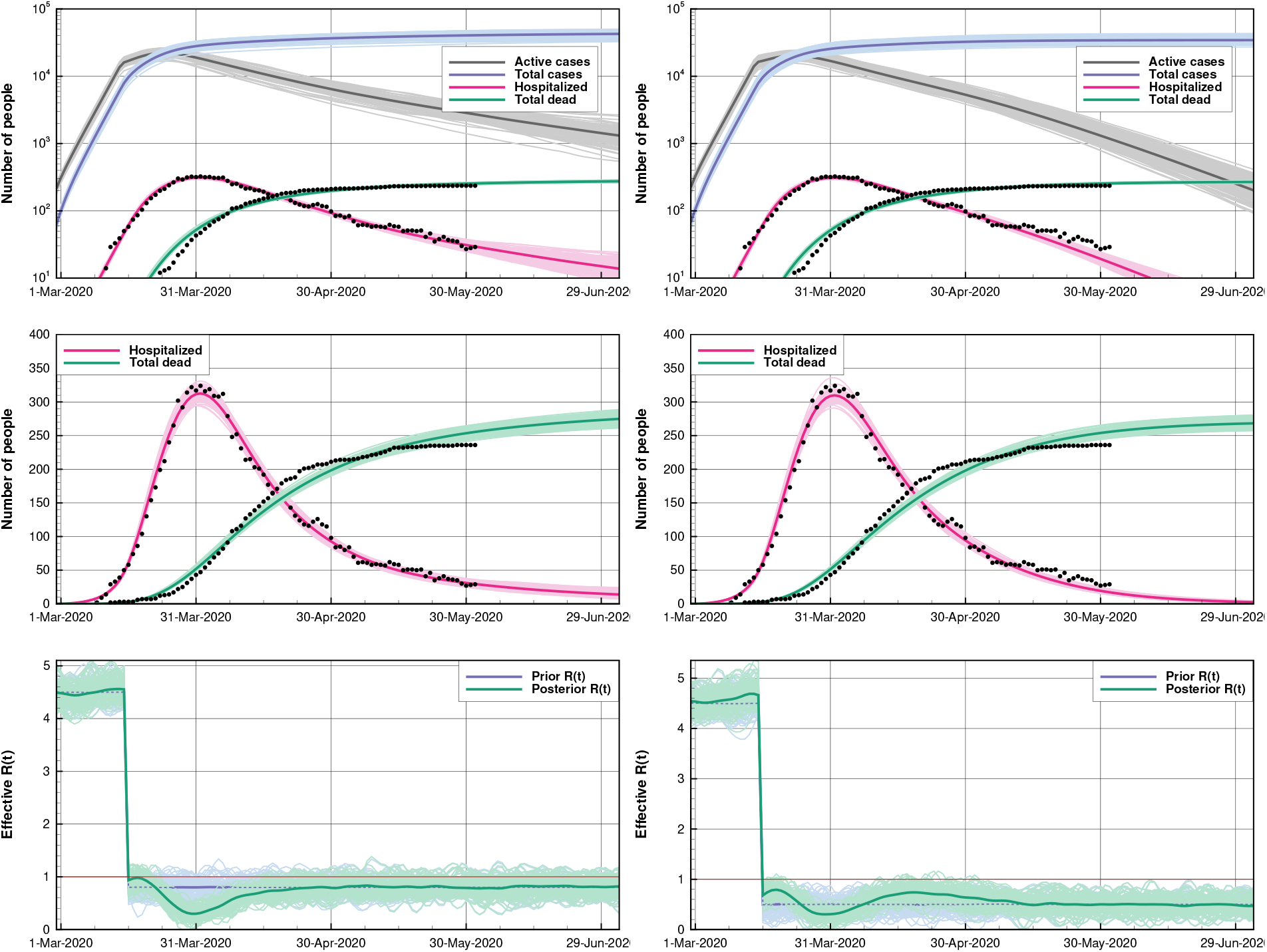
Norway (base case): For the two cases (left and right plots), the only difference is the prior-guess for *R*(*t*) after starting the interventions. For the first two rows of plots, we show the posterior ensemble means and the 100 first realizations of the posterior solution from ESMDA. The blue lines are the total number of cases, while the gray lines give the number of active cases. The red curves denote the number of hospitalized, and green lines show the total number of deaths. The upper plot uses a log *y*-axis. The second row is a zoom of the upper plot using a linear *y*-axis. The lower plots show the corresponding prior and posterior estimates of *R*(*t*) for the two cases.

These cases also confirm that the partial reopening of society led to an increase in *R*(*t*) but, luckily, *R*(*t*) stayed well below one. Also, from the current prediction, at the end of May, Norway had about 3000 active cases. If the interventions would continue to retain *R*(*t*) ∼ 0.80, then the number of active cases should be well below 1000 at the end of July, and there should only be a few hospitalized patients.

## 5. A case study for England

To date, the UK has been the worst-hit country in Europe in terms of deaths and is second only to the United States of America worldwide. Some fundamental differences between the UK and Norway help explain the differences in the evolution of the epidemic. The most important is that the UK has a much higher population density, 273 people per km^2^ compared to just 14 in Norway, with most of the population concentrated within the South-East England and London. The UK government’s response to the epidemic was also much different, initially attempting to contain and delay the outbreak and only going into lockdown once 720 deaths had already occurred in England and 11,000 people had tested positive.

Due to the different recording protocols across the devolved countries, data collection for the whole UK is not straightforward. To enable more consistent use of data, we will focus on England only, which makes up approximately 85% of the UK population.

The following chronology is useful to understand the build-up of the epidemic in England.

- By February 19th, there were a total of 20 positive cases registered. On March 5th, the first (two) COVID-19 deaths occurred.
- On March 17th, the UK Prime Minister urged people not to go to public places (pubs, theatres, gyms) and work from home. Universities started closing that week.
- On March 20th, the government passed legislation stating the closing of schools and non-essential businesses [49]. The lockdown started effectively on March 23rd.
- On May 13th, lockdown measures were slightly relieved: workers who cannot work from home have been allowed to come back to work, although they have been discouraged from using public transport.
- On June 1st, a more substantial relaxation of the lockdown measures commenced. Nurseries and schools were allowed to reopen for children up to age six and those in their final year of primary school, while bars and restaurants were to remain closed, and public gatherings were not permitted. Outdoors meetings for up to six people were allowed.

### 5.1. Data sources on COVID-19 in England

We can obtain data on accumulated deaths for England from three different sources:

- The National Health Service (NHS) England provides data on deaths recorded against the actual date of death [42]. Deaths outside hospitals, such as those in care homes, are not included.
- Public Health England (PHE) provides figures for deaths that have had a diagnosis of COVID-19 confirmed by a PHE or NHS laboratory [48]. They offer death numbers according to the day they were reported, not the day they occurred.
- Office for national statistics (ONS) publishes deaths, where the death certificate refers to COVID-19, every Tuesday [46]. These deaths include cases outside the hospital and cases where COVID-19 is suspected, but no formal diagnostic test has taken place. They only report registered deaths up to 11 days before the date of publication.

Figure 4 compares the accumulated deaths reported from the three agencies. The end dates are June 9th for the PHE (green) and NHS (blue) data. However, only data from before June 4th can be considered complete. In contrast, the ONS (orange) delay reporting of their data so that the figures will not be subject to change. The discrepancies between the three lines reflect different ways of attributing the death to COVID-19 and the date of occurrence. For instance, note how the PHE data, which reports by registration date, display a more rapid increase in deaths during the working week than at the weekends. We can obtain an estimate of the proportion of deaths occurring outside of hospitals by taking the difference of the NHS data from the ONS data. This difference increases until saturating at approximately 39% on May 29th. We, therefore, set the fraction of the fatally ill that goes in hospital as *p*_h_ = 0.7.

**Figure 4.**
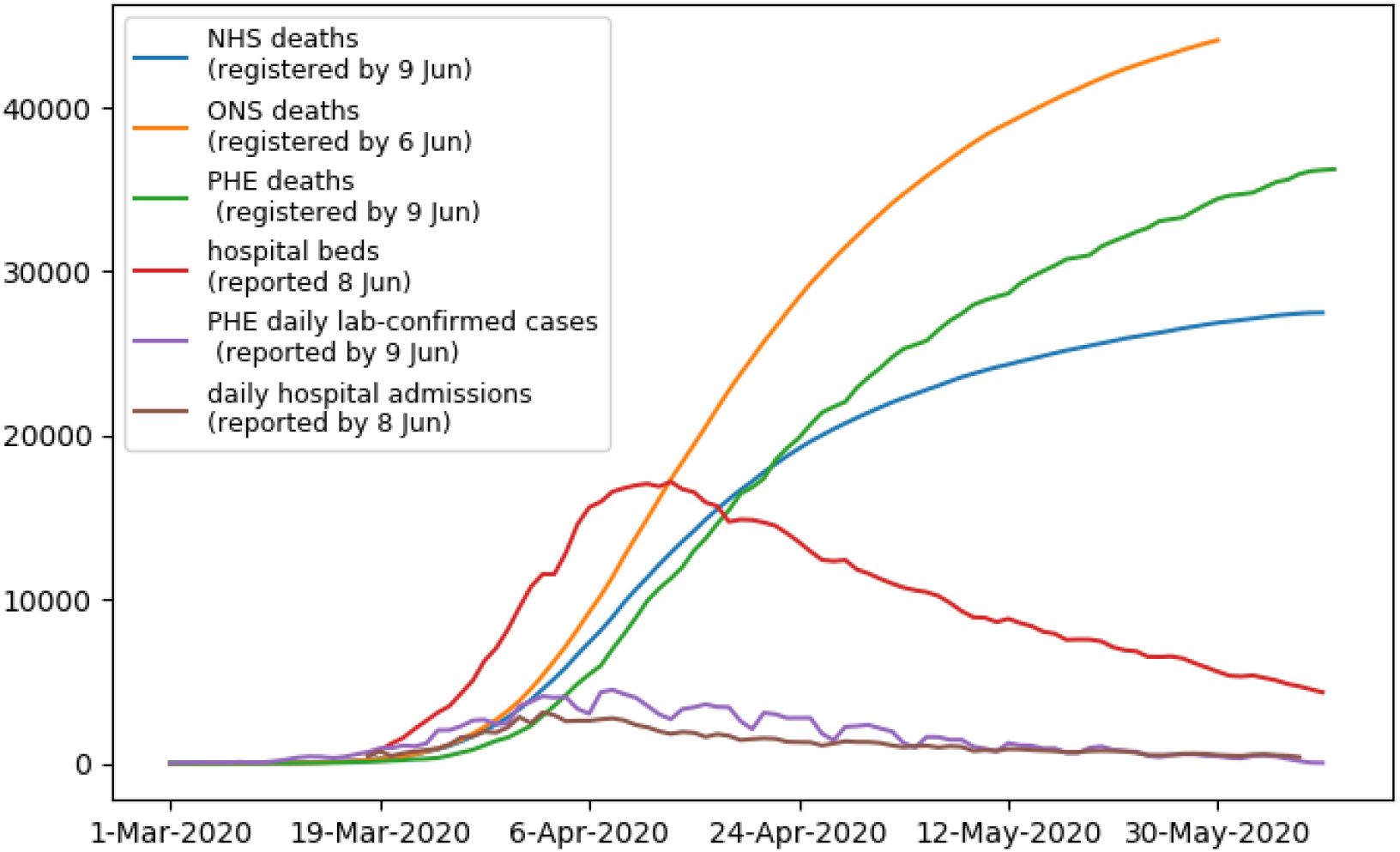
England: The plots show the data available for assimilation. Three different agencies report the number of deaths, the UK government press conference publishes the number of people in hospitals, CHESS reports the daily hospital admissions, and PHE presents the number of new cases.

The ONS data is the most appropriate for our purposes, although, due to multiple causes of death, we can not attribute the deaths solely to COVID-19. To try to disentangle this, ONS also provides data on the average number of deaths in the previous five years for comparison. For example, in the week ending April 17th, 2020, in England and Wales, there were 22,351 deaths registered, of which 8,758 attributed to COVID-19. This number compares to an average of 10,497 deaths in the previous five years for the same week. The scale of these numbers brings confidence that the new presence of COVID-19 is responsible for the dramatic increase in deaths. Unfortunately, many excess deaths are not necessarily caused by the virus but by the pressures on the NHS. Thus, more people are dying from causes, other than COVID-19, that one would otherwise cure. Our model does not cover these pandemic consequences, which pose an essential factor when interpreting the data.

Data on the number of people in hospital with COVID-19 in England and Wales is being collected by PHE and reported by the daily UK government press conferences [47]. These are the data for hospitalization used in our experiments and are plotted in Figure 4 as the red line.

For completeness, we also show the daily cases testing positive for SARS-Cov-2 reported by PHE (purple line; note that, as with the deaths, only data up to June 4th can be considered complete). It is possible to compare these cases with daily admission rates reported by the COVID-19 hospitalization in the England Surveillance System [50] (brown line). Note that the number of cases testing positive is closely following the hospital admission rates as people hospitalized have been receiving priority in testing. Hence, the number of positive cases reported can be assumed to be a significant underestimate of the true number. In Section 5.3, we will use ESMDA to quantify this underestimation and to correct the testing data before their use in the experiments.

### 5.2. Model and data assimilation setup for the case of England

We start our simulation on February 20th and assimilate data in the period from March 5th to May 29th. The model parameters were calibrated against data for England during several tuning experiments. To account for the under-sampled testing, we set the initial number of infected people to *I*_0_ = 60, three times the number of accumulated positive tests provided by PHE by February 19th. The initial number of exposed is *E*_0_ = 240. The prior value of the reproduction number before the intervention was *R*_1_ = 3.87 [29]. The model is informed about the intervention date of March 23rd by reducing the first guess for the reproduction number to *R*_2_ = 0.9, but with a relative standard deviation set to 50%. A large standard deviation accounts for a real large uncertainty of *R*(*t*) after the introduction of the intervention, but also, as seen from the results, it provides the needed variability in the ensemble members for DA. The values of the remaining parameters used for the England case are the same as for Norway (see Table 1).

The number of ensemble members (5,000), the number of ESMDA iterations (32), as well as the analysis types (stochastic EnKF) are the same as for the Norwegian experiment. Similarly, we assume observation-error standard deviations to be 5% of the data value. However, in the Norwegian example, the maximum value of the assumed error standard deviation was set to 6 people. Given the grander scale of deaths in England, we change this to be 5% of 8,000, i.e., 400. The default is that the same errors apply to the number of hospital beds occupied with COVID-19 patients as the accumulated deaths. In the experiments assimilating the number of cases testing positive, we have assumed an error standard deviation of 10% for the case observations with a maximum of 50,000. We have corrected the data to account for biases in reporting, as described in Section 5.3. As for the Norwegian example, a de-correlation half-length scale of the observations is set to 10 days to account for systematic errors in the reporting.

We have defined the pre-lockdown transmission matrix, 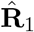, for England based on results from [40], who estimated “contact” matrices in a European study in 2006. We have adopted a transmission, 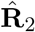, used during the lockdown, from [35], who estimated a similar “contact” matrix for the UK one day after the introduction of the lockdown. Following a normalization as described concerning Eq. (13), these two matrices provide us with the transmission matrices needed by our model to describe the inter-age group transmission before and during the lockdown. Table 4 and 5 show the two contact matrices 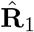 and 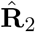. The entries of 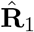 indicates that before lockdown, it is school-age children who exhibit the largest amount of contacts consistent with the school environment. Note that these matrices are not necessarily symmetric. For example, while most children will interact with adults under 50, either guardians or teachers, not all adults will necessarily have contact with children. The transmission matrix for the lockdown period, 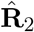, displays a large drop in the number of daily contacts. However, the reduction is not the same across all age groups, with the school-age children showing the most dramatic decrease in contacts within their age group.

**Table 4.** England: The contact matrix 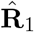 used to describe the transmission between different age groups in England before the enforced lockdown on March 23rd. The same contact matrix is used for the prediction from June 1st. See the right panel of Figure 2A in [35] for a heat-map representation of the original matrix.

| Age groups | 1 | 2 | 3 | 4 | 5 | 6 | 7 | 8 | 9 | 10 | 11 |
| --- | --- | --- | --- | --- | --- | --- | --- | --- | --- | --- | --- |
| 1 | **2.0** | 1.5 | 1.5 | 1.0 | 1.5 | 0.5 | 0.5 | 0.5 | 0.4 | 0.4 | 0.4 |
| 2 | 0.5 | **8.0** | 6.0 | 2.0 | 2.5 | 2.5 | 1.5 | 1.4 | 0.9 | 0.9 | 0.9 |
| 3 | 0.5 | 6.0 | **8.0** | 2.0 | 2.5 | 2.5 | 1.5 | 1.4 | 0.9 | 0.9 | 0.9 |
| 4 | 0.5 | 2.5 | 2.5 | **6.0** | 2.0 | 2.0 | 1.9 | 1.5 | 0.9 | 0.9 | 0.9 |
| 5 | 1.2 | 2.5 | 2.5 | 2.0 | **3.0** | 2.0 | 1.9 | 1.8 | 0.5 | 0.5 | 0.5 |
| 6 | 0.5 | 2.3 | 2.3 | 2.0 | 2.0 | **3.0** | 1.9 | 1.5 | 1.4 | 1.4 | 1.4 |
| 7 | 0.5 | 2.0 | 2.0 | 1.5 | 1.5 | 1.5 | **2.0** | 1.5 | 0.9 | 0.9 | 0.9 |
| 8 | 0.5 | 1.9 | 1.9 | 1.0 | 1.2 | 1.2 | 1.9 | **1.5** | 0.9 | 0.9 | 0.9 |
| 9 | 0.5 | 1.5 | 1.5 | 0.9 | 0.9 | 1.2 | 1.0 | 1.5 | **1.5** | 1.5 | 1.5 |
| 10 | 0.4 | 1.0 | 1.0 | 0.9 | 0.7 | 1.2 | 1.0 | 1.0 | 1.5 | **1.5** | 1.5 |
| 11 | 0.4 | 0.9 | 0.9 | 0.9 | 0.7 | 1.2 | 1.0 | 1.0 | 1.5 | 1.5 | **1.5** |

**Table 5.** England: The contact matrix 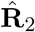 used to describe the transmission between different age groups in England during the lockdown from March 23rd to May 31st. See left hand panel of Figure 2A in [35] for a heat-map representation.

| Age groups | 1 | 2 | 3 | 4 | 5 | 6 | 7 | 8 | 9 | 10 | 11 |
| --- | --- | --- | --- | --- | --- | --- | --- | --- | --- | --- | --- |
| 1 | **1.0** | 0.9 | 0.9 | 0.8 | 1.0 | 0.5 | 0.5 | 0.4 | 0.3 | 0.3 | 0.3 |
| 2 | 0.5 | **2.0** | 1.5 | 0.9 | 1.0 | 1.0 | 0.5 | 0.4 | 0.3 | 0.3 | 0.3 |
| 3 | 0.5 | 1.5 | **2.0** | 0.9 | 1.0 | 1.0 | 0.5 | 0.4 | 0.3 | 0.3 | 0.3 |
| 4 | 0.5 | 1.0 | 1.0 | **1.2** | 1.0 | 1.0 | 0.9 | 0.5 | 0.4 | 0.3 | 0.3 |
| 5 | 0.8 | 1.0 | 1.0 | 0.9 | **1.1** | 0.9 | 0.9 | 0.5 | 0.4 | 0.3 | 0.3 |
| 6 | 0.5 | 1.0 | 1.0 | 1.0 | 1.0 | **1.1** | 0.9 | 0.5 | 0.4 | 0.3 | 0.3 |
| 7 | 0.5 | 0.6 | 0.6 | 0.9 | 0.9 | 0.9 | **1.0** | 0.7 | 0.5 | 0.5 | 0.5 |
| 8 | 0.5 | 0.6 | 0.6 | 0.8 | 0.9 | 1.0 | 1.0 | **1.0** | 0.5 | 0.5 | 0.5 |
| 9 | 0.5 | 0.6 | 0.6 | 0.6 | 0.5 | 1.0 | 0.9 | 0.9 | **1.1** | 1.1 | 1.1 |
| 10 | 0.5 | 0.6 | 0.6 | 0.6 | 0.5 | 1.0 | 0.9 | 0.9 | 1.1 | **1.1** | 1.1 |
| 11 | 0.5 | 0.6 | 0.6 | 0.6 | 0.5 | 1.0 | 0.9 | 0.9 | 1.1 | 1.1 | **1.1** |

### 5.3. Results for the case of England

We conduct three DA experiments where we assimilate different combinations of accumulated deaths (D), the daily number of hospitalized (H), and the total number of positive cases (C). The data were available for the period from March 5th to May 29th. We ran the following three experiments:

1. Exp D conditioning on D.
2. Exp DH conditioning on D and H.
3. Exp DHC conditioning on D, H, and C.

From June 1st, following the relaxation of the lockdown measures, we run predictions under three different epidemic scenarios with the prior reproduction numbers *R*_3_ = 0.5, 1.0, and 1.2. To the neutral, *R*_3_ = 1.0, scenario, we added a pessimistic one, *R*_3_ = 1.2, representing a situation without effective epidemic countermeasures, and an optimistic one, *R*_3_ = 0.5, in which effective mitigation measures are in order. For all of the prediction scenarios, we use the same transmission matrix used for the pre-lockdown period: we set 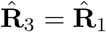.

Assimilating accumulated deaths only (top panels) brings a close fit to the corresponding data. However, the observed daily hospitalizations are substantially overestimated (cf the hospitalization data in the mid-left panel of Figure 5). The estimated effective reproductive number *R*(*t*) until the intervention on March 23rd displays a huge, possibly nonphysical, fluctuation from above 4.5 for two weeks and then decrease quickly to about 3.0 afterward. The performance of ESMDA significantly improved when we assimilated daily hospitalizations in combination with observed deaths (Exp DH, mid panels). The fit to the hospitalizations data is excellent, and the estimated *R*(*t*) at the pre-interventions period undergoes much smaller oscillations. Note, however, that its estimate for the first week after March 5th is still above the assumed basic reproduction number *R*_1_ = 3.8. After that, *R*(*t*) is below or close to 0.9 for the first half of the lockdown period. It then stays above the critical unitary value from mid-April to mid-May, and finally, slightly below 1 toward the end of the lockdown. Given that we are conditioning on data until May 29th, and the time scale of the process is two weeks, further data is required to describe the last part of the lockdown accurately.

**Figure 5.**
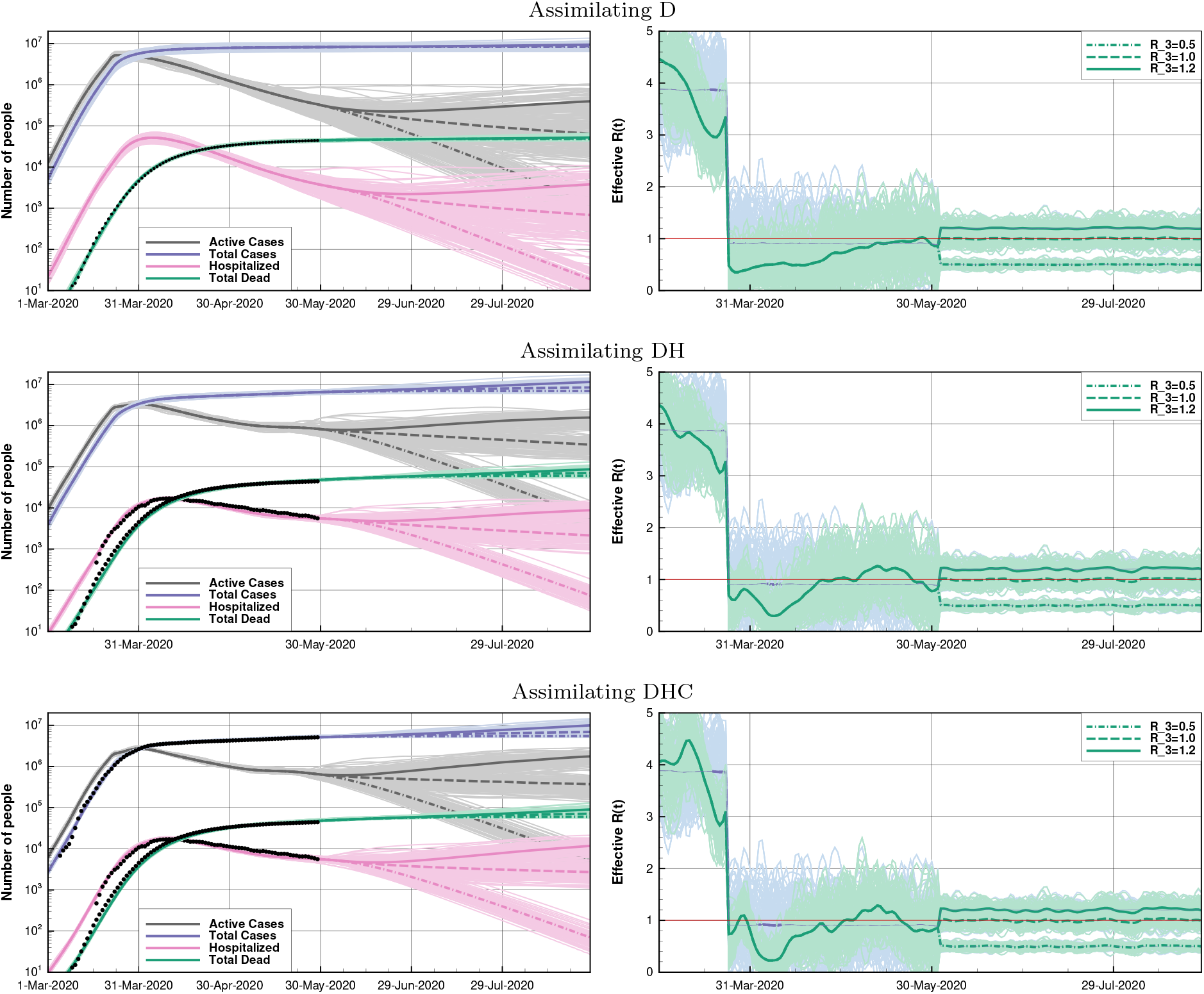
England. Left panels: ESMDA posterior estimates of the accumulated number of deaths (green), the daily number of hospitalizations (red), active (gray), and total cases (blue). Observations are displayed in black. Right panels: Prior and ESMDA posterior estimates of the effective reproduction number *R*(*t*). Ensemble members (thin lines) and ensemble means (thick lines). Assimilation experiment D (top panels), DH (mid panels) and DHC (bottom panels).

As discussed in Section 5.1, the reported number of cases testing positive for SARS-Cov-2 underestimates significantly the actual number of people infected by the virus. We can assess the magnitude of this underestimation by comparing the number of cases predicted by experiment DH to the reported number of cases. We find that as the number of people infected grew exponentially before the lockdown started and the reproduction number declined to below 1.0, the percentage of positive cases not being reported increased due to a severe lack of testing kits. On March 25th, this number peaked, with reporting of only 1% of cases. As more tests became available, and simultaneously the rate of infection reduced, the percentage of unreported cases decreased. From the beginning of May, about 2% of the accumulated cases are reported. The UK government achieved its self-imposed target of 100,000 tests per day for the first time on May 1st. The estimated percentage of reported cases can be compared to the percentage of asymptomatic cases. Estimates of the proportion of asymptomatic cases vary widely between studies. However, WHO suggests that 80% of infections are mild or asymptomatic [61], supported by [34]. Therefore, due to the testing strategy in the UK (only testing people displaying symptoms), we could conclude that approximately 10% of the accumulated symptomatic cases are being reported from the beginning of May.

To account for this change in the under-reporting of positive cases with time, we applied a piecewise correction to the data. Between March 5th and March 20th, the inflation factor needed to correct the number of positive cases reported daily increases linearly from 25 to 120. The inflation factor then decreases linearly to 12 on April 7th, where it remains until April 29th, and after that increases linearly to 60 on May 29th. We assimilated these corrected observations in the DHC experiment (shown as black points) following the total cases in Figure 5.

Given the dramatic modifications to the original data, one should assimilate these corrected observations with caution. Thus, we have increased their assumed standard deviation to 10%. Nevertheless, results in Figure 5 (bottom panels) demonstrate that the assimilation of these data leads to an additional small improvement, particularly in the fit to the (corrected) observed cases. The assimilation of these “adjusted” data for positive cases impacts the estimates of *R*(*t*) too. In particular, the results support that for about two weeks after March 5th (before the lockdown), *R*(*t*) was stably larger than 3.8. Still, it decreases to about 3.0 and finally near 1 and below during lockdown. The increase in *R*(*t*) above one from mid-April to mid-May, observed already in experiment DH, is found here too. However, as already pointed out, more recent data would be needed to constraint the solution in that period.

We present the values of the model parameters estimated by the ESMDA in the three experiments D, DH, and DHC, in Table 6, and we recall the prior values in the second column for reference. Numbers do not appear to be dramatically different between the different experiments and the priors, but we assume that the results of exp DHC are more accurate. Nevertheless, it is worth to note the differences (the increase over the prior) in *p*_s_ and *p*_f_ when assimilating hospitalizations compared to not.

**Table 6.** England: Prior and posterior mean and standard deviation for the time independent parameters estimated with ESDMA in the experiments D, DH and DHC.

| Parameters | Prior | Posterior D | Posterior DH | Posterior DHC |
| --- | --- | --- | --- | --- |
| $I_0$ | 60 (6) | 61.31 (6.01) | 61.95 (6.00) | 60.08 (5.98) |
| $E_0$ | 240 (24) | 246.69 (23.85) | 251.56 (23.62) | 239.43 (23.46) |
| $\tau_{\text{inf}}$ | 3.8 (0.5) | 2.73 (0.33) | 3.12 (0.33) | 2.83 (0.28) |
| $\tau_{\text{inc}}$ | 5.5 (0.5) | 4.61 (0.40) | 4.79 (0.40) | 5.14 (0.33) |
| $\tau_{\text{recm}}$ | 14 (0.5) | 13.99 (0.49) | 13.94 (0.49) | 13.94 (0.49) |
| $\tau_{\text{recs}}$ | 5 (0.4) | 4.98 (0.40) | 4.13 (0.31) | 3.57 (0.31) |
| $\tau_{\text{hosp}}$ | 6 (0.5) | 5.54 (0.49) | 5.35 (0.48) | 4.79 (0.39) |
| $\tau_{\text{death}}$ | 16 (0.5) | 15.62 (0.50) | 15.12 (0.47) | 14.47 (0.44) |
| $p_f$ | 0.009 (0.001) | 0.009 (0.0009) | 0.011 (0.001) | 0.015 (0.0001) |
| $p_s$ | 0.039 (0.003) | 0.039 (0.003) | 0.011 (0.001) | 0.015 (0.001) |

From June 1st to September 1st, we run the three scenario-predictions for each of the assimilation experiments D, DH, and DHC. For the optimistic case, *R*_3_ = 0.5 (dot-dashed line), the differences in the forecasts between the cases D, DH, and DHC, are minor. However, case D tends to differ more (in particular to predict substantially fewer deaths and hospitalizations), arguably as a consequence of not being constrained on hospitalization data. Even in this more favorable scenario where we assume the measures will keep *R*(*t*) as small as 0.5, experiments DH and DHC predict more than 60,000 casualties by September 1st. Note that experiment D predicts only 48,000 dead. Thus, even in this “favorable” scenario, the capability of simultaneously assimilating multivariate observations (deaths, hospitalization, and cases) allows for tracking 10,000 more deaths that we could potentially have saved.

For the neutral case, *R*_3_ = 1.0 (dashed line), DH and DHC predict 70,000 deaths and 2,300 hospitalization, while D predicts “only” 50,000 deaths and 720 hospitalizations by September 1st. Further differences between the three experiments become evident when *R*_3_ = 1.2 (solid line). Experiments DHC and DH predict 90,000 deaths, against 50,000 for D, and 10,000 hospitalizations against less than 5,000 for experiment D. Note that DHC predicts systematically for all three values of *R*_3_ considered, slightly more deaths and hospitalizations than DH. Given the possible human drama behind each digit of increase, the use of the three, albeit biased, data-sets may have an enormous social impact. This example confirms the importance of having access to a variety of useful data and of their multivariate treatment that data assimilation offers. Still, they, unfortunately, picture a very gloomy future.

## 6. A case study for Quebec, Canada

Data for Quebec are available from the INSPQ [3], which is the national public health institute for Quebec. Data until May 28 have been used (the data were extracted on May 29). These include the accumulated number of deaths, the number of positive cases, and the current number of hospitalizations. The fatalities also include those occurring in care homes for the elderly and any situation where COVID-19 was a direct contributing factor. It does not include any deaths due to other causes such as delayed medical interventions in a stressed health care system or fear by sick persons of getting infected during a necessary hospital visit. The deaths can be attributed either to the day on which they were reported or the day on which they occurred. Counts for a specific day of occurrence become relatively stable after about seven days. However, changes to the database will continue to be made as appropriate.

In addition to reported hospitalizations and positive tests, we condition on the reported deaths *d*_R_ to estimate the model parameters. To reflect the approximately four-day delay between an actual or modeled death, *d*(*t*_*i*_), at day *t*_*i*_, and the reported death, we use:

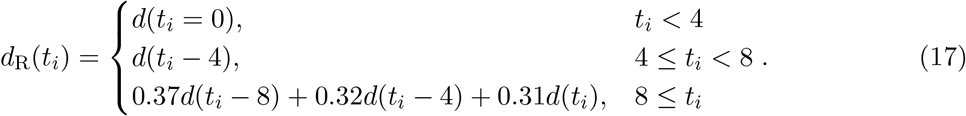

We applied this equation to the model predicted deaths, *d*(*t*_*i*_), whenever comparing the model values and the observations, *d*_R_(*t*_*i*_), such as for a figure.

An estimate of the excess mortality during the epidemic can be obtained from weekly data published by the Quebec Statistical Institute (ISQ) [2]. As of May 15th, the weekly totals were available until April 25th. When computing the accumulated mortality for the period of March 1st until April 25th, for each of the five years from 2015 until 2019, the average excess amounts to 1919 deaths. The standard deviation in the five individual values is 218. For April 25th, the INSPQ reports a total of 1921 occurred deaths, which is remarkably close to the value provided by the ISQ. However, it is noted by the ISQ that the estimate of the most recent weekly total deaths only covers about 80% of the real number. Thus some evidence of additional excess mortality related to COVID-19 might appear in the future. It is also noted that for the same date of April 25th, data were extracted from both sources on May 29th. The ISQ data indicated an excess of 2019 deaths, and the INSPQ total was 2001, which was still in good agreement.

The number of hospitalizations is possibly accurate, but the fraction of fatally ill that goes to the hospital was not known for this study. A large number of deaths occurred at care homes for the elderly, so we used a fraction, *p*_h_ = 0.5, in the experiments. On May 20th, the number of hospitalizations was reduced by approximately 200, due to a different way of counting. From May 20th onward, recovered patients still in the hospital awaiting a transfer were removed from the number of hospitalized patients. We distributed this shift of 200 over the preceding days as a sequence of 40 per day. This approach avoids a jump in the assimilated data.

The number of positive test cases includes likely associated cases, such as family members, where testing was deemed unnecessary. Initially, testing had to be limited to the subjects that were most likely to be positive, notably travelers, and, as the number of available tests improved, more groups were tested. Thus, over time, the testing became more comprehensive. Over the entire period, the assumption is that only 15% of positive cases show up in the reported statistic. Thus the reported number of cases was multiplied with a factor of 1/0.15 for use in the assimilation experiments. For the three data types, we use a relative error of five percent. We have neglected possible delays in the testing and reporting procedures for the number of positive cases. Finally, we obtained the population data from the Quebec Statistical Institute for 2019 [1]. The total population is 8,484,965 and, as before for Norway, available per age range.

Table 7 gives the parameters used for the epidemic in Quebec. The prior value for the case fatality rate is, with a value of 0.02, rather high, which reflects that the epidemic hit the most fragile part of the population very hard. The uncertainty in the parameter values is about twice what is used for the other cases discussed in this manuscript. We choose these larger values to have an appropriate amount of uncertainty in the forecast ensembles. Contributing factors in the early development of the epidemic were the “Spring Break,” which was from March 1st until March 6th, as well as close connections between the metropolitan areas of Montreal and New York. The SEIR model does not capture such events. Instead, the model is starting with relatively high estimates of the initially exposed and infectious. Although restrictive measures were introduced gradually over several days, we assume that they all became effective on March 23rd. For the initial period of free exponential growth, we use a prior estimate of *R*_1_ = 3.0 ± 0.6 for the effective reproductive number. To not bias our posterior estimate of the effective reproduction number to values above or below unity, the prior estimate is *R*_2_ = 1.0 ± 0.5 for the second (and final) period. We estimate the effective reproductive number until May 28th and keep it constant after that. Note that the available data only weakly constrain the effective reproductive number in the final week.

**Table 7.** Quebec: The set of prior model parameters and their standard deviations (a zero std dev denotes that the parameter is kept fixed). Columns DHC, DH and D show posterior values for, respectively, experiments DHC, DH and D. Note that *p*_*h*_ was supplied externally. The curves for *R*_1_ and *R*_2_ are shown in Figure 6.

| Parameters | Prior(Std Dev) | DHC | DH | D |
| --- | --- | --- | --- | --- |
| $R_1$ | 3.0(0.6) | - | - | - |
| $R_2$ | 1.0(0.5) | - | - | - |
| $I_0$ | 100.0(20.0) | 67 | 98 | 96 |
| $E_0$ | 240.0(48.0) | 167 | 204 | 235 |
| $\tau_{\text{inc}}$ | 5.5(1.0) | 5.2 | 3.4 | 3.8 |
| $\tau_{\text{inf}}$ | 3.8(0.6) | 1.8 | 1.9 | 2.7 |
| $\tau_{\text{recm}}$ | 14.0(2.0) | 14.1 | 12.8 | 14.8 |
| $\tau_{\text{recs}}$ | 5.0(1.0) | 6.9 | 6.8 | 5.5 |
| $\tau_{\text{hosp}}$ | 6.0(1.2) | 5.9 | 5.8 | 6.7 |
| $\tau_{\text{death}}$ | 10.0(2.0) | 5.8 | 3.4 | 10.4 |
| $p_f$ | 0.020(0.004) | 0.020 | 0.021 | 0.023 |
| $p_s$ | 0.039(0.006) | 0.040 | 0.047 | 0.038 |
| $p_h$ | 0.5(0) | - | - | - |

### 6.1. Data assimilation experiments

Similar to the case for England, we have performed three types of experiments: DHC - assimilating deaths, hospitalizations, and positive cases; DH - assimilating deaths and hospitalizations, and D assimilating deaths only. The upper plots of Figure 6 shows the results for experiment DHC. The actual and reported number of fatalities match rather closely from the beginning of the epidemic. For the hospitalizations, the curves match after the initial week and for the positive cases (not shown) after the initial two weeks.

**Figure 6.**
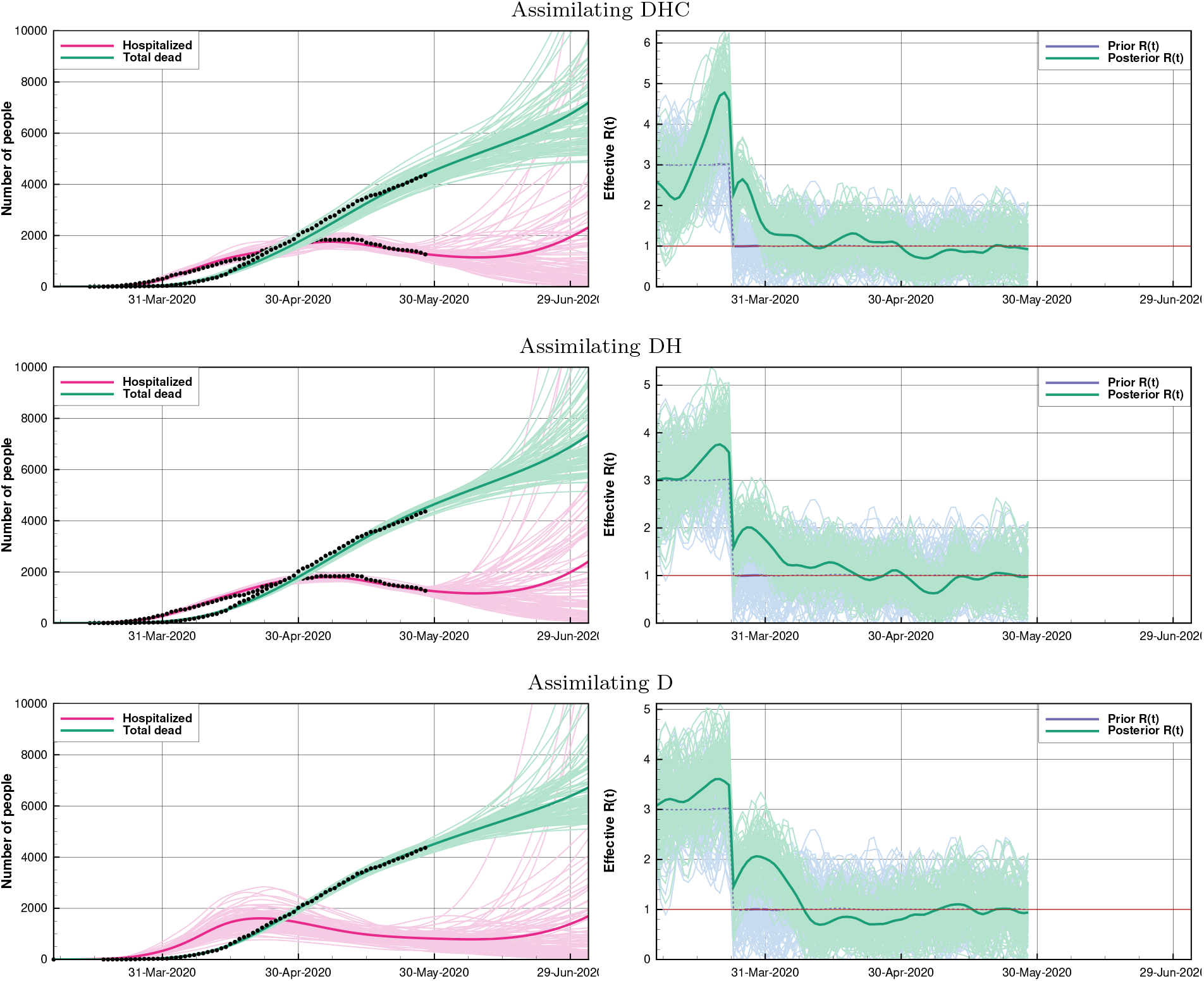
Quebec: From top to bottom, the plots show the results from the three assimilation experiments DHC, DH, and D. The left column presents the accumulated number of deaths and the number of hospitalizations, and the right column shows corresponding the reproductive number *R*(*t*) for the experiments. Time is since the start of the epidemic on March 8th. Observations are indicated with points when used to obtain the model fit. The solid lines are for the ensemble mean posterior estimates. After May 28th, we kept the realizations of *R*(*t*) constant and equal to the latest values for the remainder of the simulation.

We note that after the initial period of rapid growth in March, the epidemic gradually slowed down in April. As of May, the numbers of new cases, new deaths and hospitalizations are relatively constant, or slowly decaying, resulting in near-unity estimates of the reproductive number. The gradual reduction in the reproductive number may reflect both the gradual introduction of more restrictive measures for the general population and the progressive improvement of procedures in care homes to control the epidemic.

The middle plots of Figure 6 show the results for experiment DH. The numbers for the initial infectious and exposed are higher than in the experiment DHC (Table 7). Note, however, that the fraction of detected cases, which was assumed to be at 15% for experiment DHC, is essentially an unknown variable. Thus, additional information on the percentage of infected people would be necessary to calibrate the fraction and permit to make a direct connection between the numbers of cases and the numbers of infectious and exposed cases.

Finally, the lower plots of Figure 6 shows experiment D, in which we only assimilated the observed number of deaths. All experiments feature a reasonably good fit between modeled and observed values, with generally minor differences. The reduction in hospitalizations in May suggests that the epidemic has peaked for all experiments. With experiments DHC and DH, we do not fit the reduced rate of new deaths in May as well as with experiment D that only assimilated this data. This issue reflects the difficulty of having coherence between the SEIR model and all observed variables over the entire experimental period. Note that experiment D, which did not use the information on hospitalizations, features a broad spread of the ensemble for this variable. The increase of the mean-predicted number of hospital admissions in the last weeks of June is a consequence of having a few members with rapid exponential growth.

For the three experiments, the first wave of the epidemic likely peaked between mid-April and early May and gradually began to decline. In June, due to the exponential nature of the outbreak, the uncertainty grows rapidly during the forecast. The gradual opening up of society in June would ideally be supported by recent data on the evolution of the epidemic. It is thus essential that these data become available more quickly. Concerning predictions for a possible second wave of the outbreak and the eventual arrival at group immunity, it will be essential to perform additional random tests to estimate the fraction of infected persons.

### 6.2. Predictability experiment

As a verification of the general methodology, we performed a sequence of retrospective 2-week forecasts for the configurations DHC, DH, and D. The first of these forecasts use data until April 1st and are valid on April 15th. Successive projections are separated by one week, and the final predictions were issued on May 13th and valid for May 27th. The results in Figure 7 show that the size of the forecast errors corresponds reasonably well with the plus or minus one standard deviation range of the ensemble of model forecasts. Initially, the deaths are underpredicted, while during the last part, they are overpredicted.

**Figure 7.**
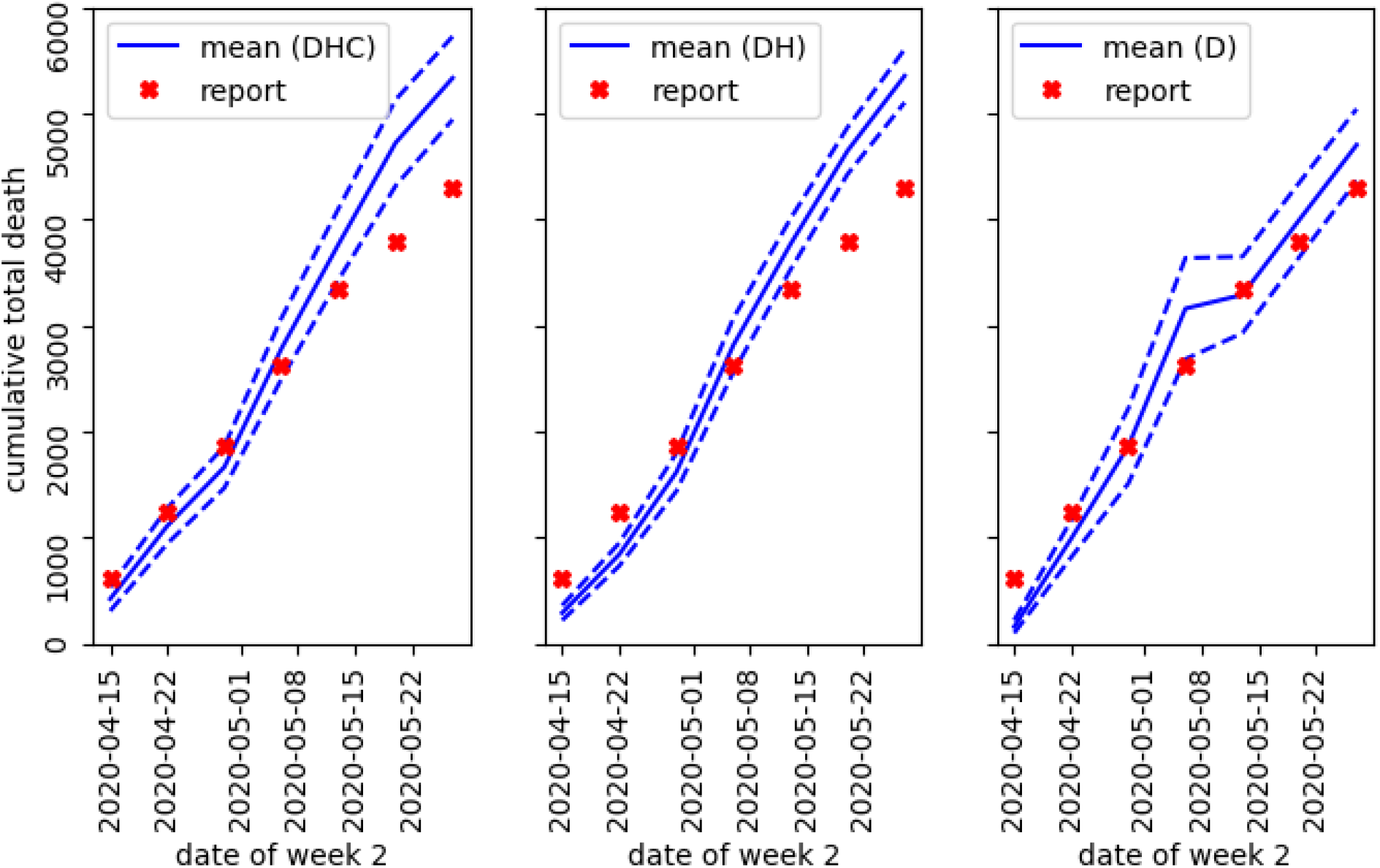
Quebec: The plots show verification of retroactive week-two forecasts for experiments DHC, DH, and D. For the predictions, issued one week apart, we show the mean estimate with the full line, and the dashed lines give the mean value plus or minus one standard deviation. We indicate the reported values with crosses.

The unsuccessful forecast issued on April 1st, with configuration D, is shown in the left plot in Figure 8. Despite restrictive measures being in place, the virus started super spreading in houses for the elderly, leading to a rapid increase in the reported number of deaths as early as April 2nd. By April 15th, the actual number of fatalities is beyond the extreme members of the 1000 member forecast ensemble. We obtain a much better forecast with the DHC configuration (right plot of Figure 8). As positive cases and hospitalizations typically precede deaths by some days, the assimilation of these data help for the prediction of the rising number of deaths. The experiments DHC and DH do, however, overpredict the number of reported deaths for the last two forecasts in May (Figure 7). A hypothesis is that the improving procedures in the health care system, notably in the care homes, made the COVID-19 epidemic less fatal. Also, additional testing and contact tracing likely increased the fraction of detected asymptomatic cases, which would lead to an overprediction of deaths. Finally, the apparent improvement may be related to reporting delays. On May 30th, an unusually large number of 202 deaths, was reported in an end-of-month adjustment.

**Figure 8.**
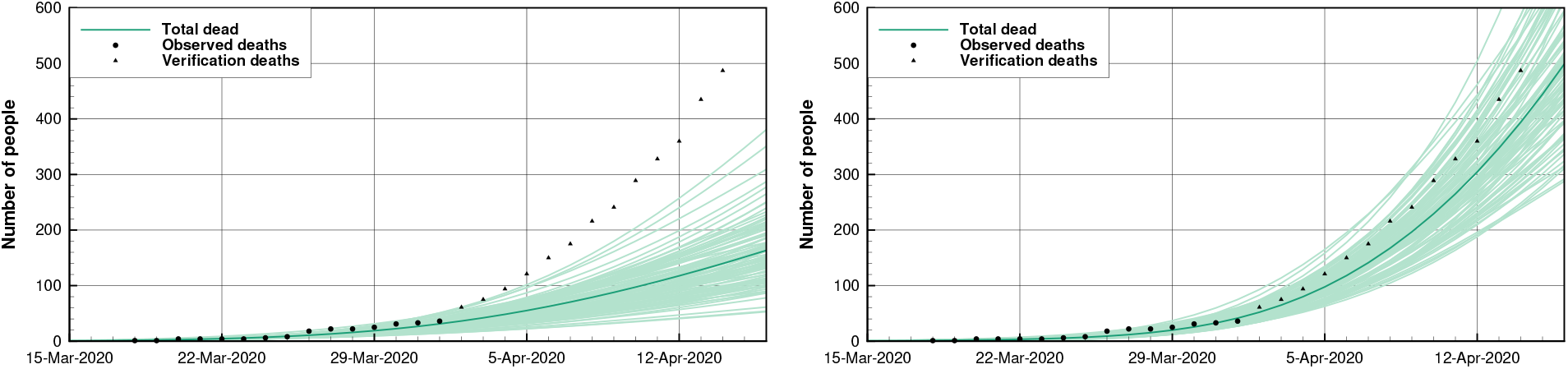
Quebec: The plots present verification of the week-two forecasts (retroactively) issued on April 1st for experiment D (left) and the experiment DHC (right). The solid line denotes the mean prediction. The reported values used to fit the model parameters are indicated with circles, while the triangles are the values used for verification.

The need for unexpectedly large standard deviations, for the prior parameter values, may reflect that the SEIR model cannot describe the full complexity of the evolution of the epidemic in Quebec. Because of the general progression in response to the outbreak, it could be appropriate to tune model parameters separately for the different phases of the epidemic.

## 7. A case study for The Netherlands

We have applied an SEIR model with the ensemble data-assimilation method to study the effect of the prior assumptions on *R*(*t*) on the estimates of the spreading of COVID-19 in The Netherlands. The first observed COVID-19 infection occurred on February 27th in the province of North Brabant. On March 1st, the government advised anyone with COVID-like symptoms in this province to stay home. On March 9th, the inhabitants of North Brabant were encouraged to work from home as much as possible. Three days later, the rest of the country followed this advice. When schools and daycare centers and restaurants and bars were closed down on March 16th, the contact between people decreased further. Children under the age of 12 years old were still allowed to play outside. Thus, the interaction among children of this age group has only slightly decreased due to government measures.

The spreading of the virus increased the pressure on the Dutch healthcare system, with a peak of 611 patients admitted to hospital on a single day on March 27th [56]. The number of registered infections per day reached a maximum of 1398 on April 10th [56]. On April 21st, when the number of new infections and hospitalizations showed a consistent decrease, the Dutch government announced a slightly less stringent lockdown with the possibility for children until the age of 18 to practice outdoor sports as of April 28th, and the re-opening of daycare centers and elementary schools for children until the age of 12 as of May 11th. By this time, also outdoor sports for groups of adults became possible, albeit at 1.5 m distance. A further opening-up took place on June 2nd, with restaurants and bars opening. In the same week, high schools re-opened while respecting the 1.5 m distance between students. Throughout Dutch society, the population adheres to the imposed 1.5 m distance between individuals, except for those using public transportation.

Data on registered infections, hospitalizations, and deaths as a result of the COVID-19 virus are shared daily by the Dutch RIVM [56] and in the Dutch media. The number of COVID-19 patients, present in intensive care units (ICU), has been monitored intensively by the Dutch foundation of national intensive care evaluation (NICE), which also publishes numbers on the total number of COVID-19 patients in hospitals [44, 45]. For this manuscript, we use the hospitalized and ICU patient numbers from NICE and the death count from RIVM; all downloaded on June 3rd, 2020. For the partitioning of the population between the different age groups, we use the data of Statistics Netherlands (CBS) [15]. The reproduction number itself is assumed to be independent of age, so the distribution of *p* numbers across ages is a direct function of the populations of the different age groups, similar to the Norwegian *p* numbers presented in Table 2.

We observe a strong linear correlation between the number of ICU patients with COVID-19 and the number of hospitalized COVID-19 patients during the onset of the epidemic. This relationship led us to conduct experiments in which we assimilated the numbers of ICU patients rather than the more uncertain data on the number of hospitalized patients. The correlation changes after March 27th, shortly before the number of ICU patients reaches a maximum of 1313 on April 7th. When the number of ICU patients is consistently decreasing, we observe a different correlation. This effect is likely related to the difference in time that patients stay in ICU and regular care units, and to the transition of patients between both departments. This variability in the correlation between the numbers of ICU patients and hospitalized may have consequences for the suitability of the observed numbers of ICU patients in a model that is tuned to assimilate the total number of hospitalized.

We consider four different cases. In all cases, we assimilate the number of deaths, the number of ICU patients (Case 1DI), or the number of hospitalized (Cases 1DH, 2DH, and 3DH); see Table 9. In Cases 1DH and 1DI, the prior reproductive number *R*_1_ = 3.8 before any country-wide intervention. After that, we reduced it to *R*_2_ = 0.8. In Case 2DH, *R*(*t*) = 1.0, while in Case 3DH, *R*(*t*) is a function of time that starts at 1.8 and gradually reduces to 0.8. Out of these four cases, the prior *R*(*t*) of Case 3DH resembles the gradually changing *R*(*t*) shared by the RIVM the most.

Experiments that assimilate the number of infected struggle to fit all three data types. Given that the number of registered infected may not be representative of the total number of positive cases, we decide not to use these data.

We have performed a large number of sensitivity-runs to evaluate the choice of prior parameters for The Netherlands (not shown). Based on this, we have decided to use the first-guess values given in Table 1. In Table 8, we provide an overview of the country-dependent parameters used in the simulations.

**Table 8.** The Netherlands: The table gives values of the parameters used in Case 1, for the parameters that are different from those indicated in Table 1. The starting date of the simulations is February 20th, 2020.

| Parameter | First guess | Std. Dev. | Description |
| --- | --- | --- | --- |
| $E_0$ | 500.0 | 50.0 | Initially exposed |
| $I_0$ | 400.0 | 40.0 | Initially infectious |
| $R_1$ | 3.8 | 0.05 | Reproduction number before interventions (Case 1DH and Case 1DI) |
| $R_1$ | 0.8 | 0.01 | Reproduction number after first nation-wide intervention (Case 1DH and Case 1DI) |
| $R_1$ | 0.95 | 0.75 | Reproduction number (Case 2DH) |
| $p_s$ | 0.010 | 0.0001 | Hospitalization rate (Case 1DI) |
| $p_s$ | 0.039 | 0.0039 | Hospitalization rate (Case 1DH, Case 2DH and Case 3DH) |
| $p_h$ | 0.5 | | Fraction of fatally ill going to hospital (Case 1DI) |
| $p_h$ | 0.6 | | Fraction of fatally ill going to hospital (Case 1DH, Case 2DH and Case 3DH) |

**Table 9.** The Netherlands: overview of cases.

| Name | Assimilated data | Description |
| --- | --- | --- |
| Case 1DI | deaths, ICU patients | prior $R(t)$ equals 3.8 before intervention, 0.8 after |
| Case 1DH | deaths, hospitalized | prior $R(t)$ equals 3.8 before intervention, 0.8 after |
| Case 2DH | deaths, hospitalized | prior $R(t)$ equals 1.0 |
| Case 3DH | deaths, hospitalized | prior $R(t)$ equals 1.8 at start of simulation and gradually ramps down to 0.8 |

In Case 1DI, in which we assimilate ICU patients rather than hospitalized patients, the use of a lower value of *p*_s_ reflects the difference in hospitalization rate between these two groups of patients. The numbers set for *I*_0_ and *E*_0_ for the initial infected and exposed are rather high compared to the number of registered positive cases. This choice accounts for the fact that the number of registered infected is likely an order smaller than the actual number of infected.

The number of deaths and hospitalized or ICU patients for Case 1DH or Case 1DI, respectively, are shown in the two top panels on the left-hand side of Figure 9. In Case 1DI, the estimated number of ICU patients has a reasonably good fit to the data. The fit to the observed number of hospitalized in Case 1DH is poorer, especially during the onset of the epidemic. The uncertainty of these numbers possibly caused this more significant misfit. In all simulations, the estimated number of deaths tends to be lower than the observed number of deaths in the onset of the epidemic. It then increases more strongly in the last days of the assimilation period.

**Figure 9.**
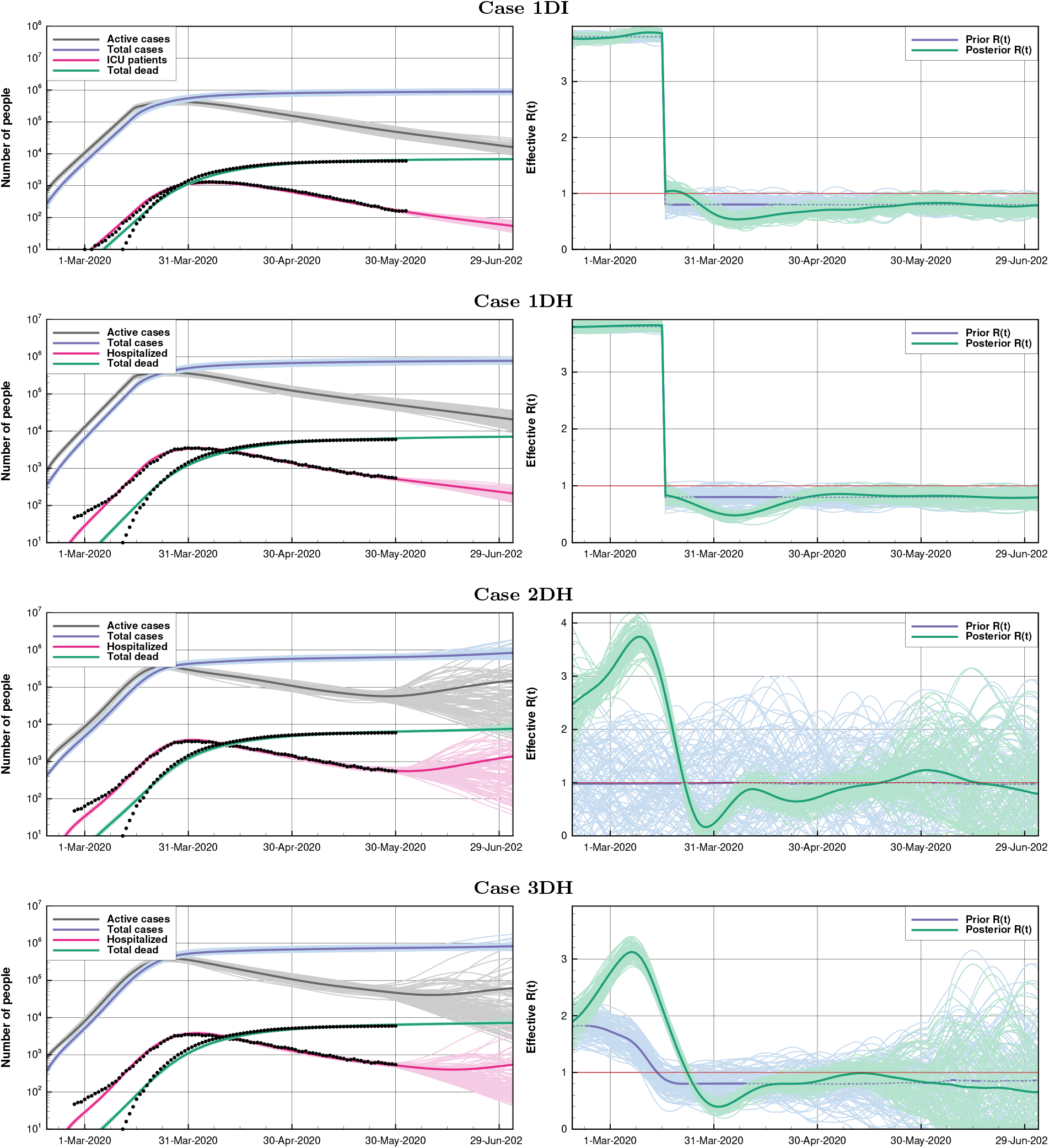
The Netherlands: The plots show the results from the Cases 1I, 1H, 2H, and 3H, from top to bottom. The left plots include model estimates of the number of hospitalized patients and dead, in addition to the total number of cases as well as the number of active cases. The right plots show the corresponding estimates of *R*(*t*).

The observations suggest a stabilization of COVID-19-related deaths of slightly over 6000, while our simulations indicate a total of approximately 8000 deaths. Because of the logarithmic scale, this difference is not clear in Figure 9. Statistics Netherlands (CBS) reported a substantial number of excess deaths in late March and early April [16]. The total number of excess deaths, based on weekly estimates, adds up to 8765 over March 9th to May 10th. Based on available data for the years 2015-2019, we estimate the standard deviation of reported deaths in this period to be around 670. The number of reported COVID-19-related deaths over this period is 5597. RIVM suggests a correlation between the epidemic and the higher number of excess deaths in early April and states that the actual number of people dying is higher because not all people who die in the Netherlands have been tested for COVID-19 [55]. The under-reporting of the number of COVID-19 related deaths could explain why our estimates for the total number of deaths are higher than what the number of reported deaths would suggest. As a consequence, our relatively high estimate of the number of deaths might be considered a rough estimate of the actual number.

In both the Cases 1DI and 1DH, the posterior estimate of *R*(*t*), as depicted in Figure 9, appears to fit the prior value of 3.8 relatively well. The dip in *R*(*t*) after the start of the first intervention, should be interpreted as an adaptation effect to the sudden change in the prior *R*(*t*). The differences between estimates of the number of deaths and *R*(*t*) between the Cases 1DI and 1DH are relatively minor. For consistency with the other experiments in this paper, the additional cases shown, assimilate the numbers of hospitalized patients.

To investigate the evolution of *R*(*t*) without relying on the prior estimate, Case 2DH assumes a constant first-guess value of 0.95 and a relatively high standard deviation of 0.75. The resulting fit to the data is comparable to Cases 1DI and 1DH. The evolution of *R*(*t*) for Case 2DH, which is illustrated in Figure 9, is different from Cases 1DI and 1DH. Because of the constant prior *R*(*t*), the posterior *R*(*t*) tends to change gradually. When the actual *R*(*t*) changes more rapidly, the model estimate will respond more slowly and exhibits a compensation of an *R*(*t*) that drops below 0.5. The conditioning of the data in Case 2DH results in a peak of *R*(*t*) around 3.8, roughly corresponding to the first guess for the initial R in Cases 1DI and 1DH. The effectiveness of the interventions in reducing *R*(*t*) is visible in the *R*(*t*) staying below or close to the value of 1 from late March onward.

Case 3DH uses a prior *R*(*t*) that mimics the estimate of *R*(*t*) published by the RIVM[54]. Rather than having a step-wise change of *R*(*t*), it assumes a gradual decrease from 1.8 to 0.8 over the first couple of weeks of the epidemic. Thus, it reflects an incremental impact of the measures. In this run, we allow the *R*(*t*) to be influenced by the data assimilation by imposing a more significant uncertainty, although smaller than in Case 2DH. The resulting estimates of dead and intensive care patients are relatively accurate. The posterior *R*(*t*) increases rapidly in the first weeks, suggesting that the prior estimate of an initial *R*(*t*) was possibly too optimistic. See Figure 9 for the results.

The results with the different prior assumptions for *R*(*t*) illustrate that we can use different priors for *R*(*t*) and still fit the data. Estimates of *R*(*t*) tend towards values around three in the onset and just below one after the peak of the epidemic. Slight changes of short duration in *R*(*t*) do not appear to have much effect on the pandemic’s evolution. The data-assimilation system appears to favor a gradual development of *R*(*t*) and our results suggest that compensation effects occur when the actual *R*(*t*) is less smooth.

We have simulated additional cases where *R*(*t*) increases to 1.0 at the moment when the schools re-opened in May. In these simulations, the number of COVID-19 deaths continues to grow and does not stabilize as it would do in Cases 1DI and 1DH (Figure not shown). We can conclude from this that for the case of The Netherlands, continued vigilance is of crucial importance. These results also make clear that further intensification of contact between the Dutch population has the potential to result in an unstable growth of the number of COVID-19-related deaths.

## 8. A case study for France

We have applied the SEIR-based ESMDA data assimilation method with data from the pandemics in France to assess the effect of the lockdown and to run possible scenarios following the intervention.

The first occurrences of confirmed cases of COVID-19 in France were reported at the end of January 2020. However, biological a posteriori sample testing showed that there were isolated cases as early as November 2019. The first clusters of infectious patients were identified at the end of February in the Oise district (north of Paris area) and in the city of Mulhouse (east of France) at the beginning of March. Stage 3 of public intervention, which recognizes the pandemics, started March 14th with the closure of schools and universities, and the enforcement of the first systematic restrictions on public meetings. A full lockdown was enacted on March 17th. The primary goal of this lockdown was to avoid the saturation of intensive care units (ICUs) in hospitals (whose theoretical capacity was 4000 beds). Occupied ICU beds rose to 7,148 on April 8th before decreasing. Nonessential business and, to some extent, schools progressively re-opened on May 11th while enforcing physical distancing. In stage 2 of the lockdown-ending, restaurants, bars, and indoor sports facilities re-opened on June 2nd, together with lifting the restrictions on traveling beyond 100km, with a few exceptions for Paris region where hot spots (such as in the Val d’Oise district) might still have a reproduction number higher than one.

Santé Publique France provides the data for France and makes them available on their daily updated dashboard [30]. These data have been officially published since March 2nd (see Figure 10). The published data are the cumulative deaths at the hospitals, the current number of hospitalized patients for confirmed COVID-19 cases, and the current number of (COVID-19 related) patients in intensive care units. These numbers apply to the whole country. Although regional data are also available, we do not exploit them here. Also, the daily number of patients released from the hospitals, and the total number of confirmed cases are provided. From mid-March, the dashboard also offered the number of deaths in care homes and other medical institutions (except hospitals) and confirmed cases in these institutions. Given the lack of reactive compounds, PCR (polymerase chain reaction) testing was only applied to suspected patients and was initially systematically carried out in the hospitals. Hence the number of confirmed cases is by far not representative of the actual number of infected people.

**Figure 10.**
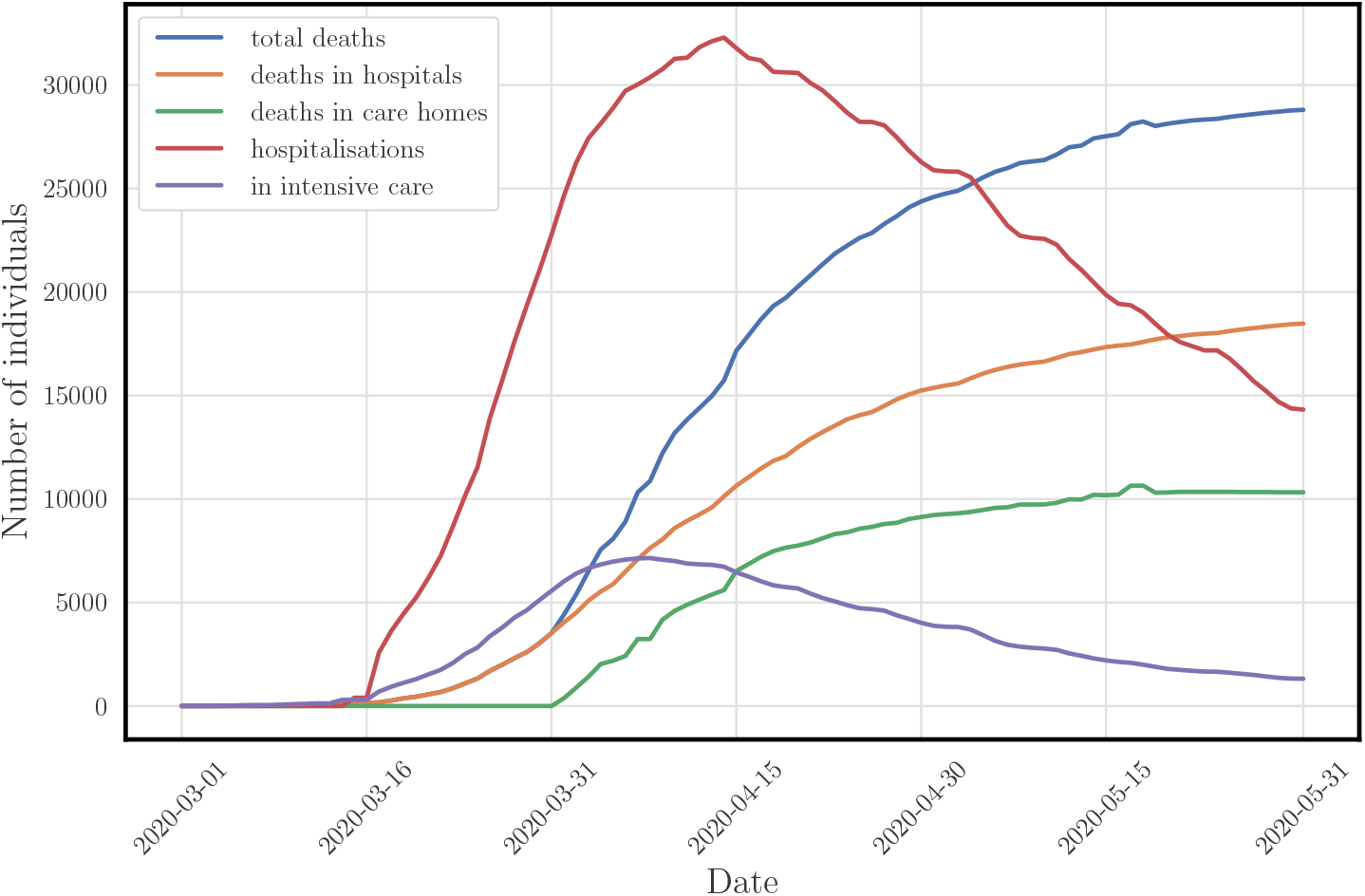
France: The plot shows official data curves for France from SantéPublique France up to May, 31.

Inaccuracies and delays in reporting of deaths in care homes to SantéPublique France profoundly impact the accuracy of the total deaths observations. Moreover, we believe that, at least for France, we cannot model the deaths in care homes with the same SEIR-like dynamics as we do for the rest of the country. Care homes comprise confined spaces with an aged, weakened population with a very different 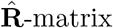 than the rest of the country. Thus we do not use these data in the conditioning. By contrast, the numbers of deaths at hospitals are likely more reliable.

### 8.1. First experiment: Calibrated reference

The first experiment serves as a reference for the application of ESMDA on the data for France. We have chosen the model parameters based on several sensitivity simulations, which yield a calibrated reference analysis. First of all, only the accumulated deaths at hospitals, together with the hospitalized, are assimilated. We have also experimented with the total deaths, which include the deaths in care homes. However, as explained above, we do not believe that, at least for the French data, they can easily be modeled by a global SEIR model. By contrast, the deaths at hospitals as well as hospitalized are expected to be reliable and representative so that we choose a standard observation deviation of 1%, with a maximum error of 50 individuals. The experiments start on February 16th and end by late August.

The reproduction numbers before the intervention and during lockdown are following INSERM (Institut national de la santé et de la recherche médicale), and Institut Pasteur studies [57]: *R*_1_ = 3.5 with a standard deviation of 0.2 and *R*_2_ = 0.65 with a standard deviation of 0.2. We also choose *R*_3_ = 0.77 with a standard deviation of 0.2 for the reproduction number after the second intervention (re-opening). This latter reproduction number corresponds to a preliminary estimate from the French government on May 28th. Moreover we chose *τ*_recs_ = 20 days, *τ*_death_ = 7 days and *p*_f_ = 0.02 as it seems to better reflect the situation in French hospitals. We set the initially exposed and infectious set to somewhat arbitrary values but with a huge standard deviation to account for the large uncertainty. Other key parameters have been left unchanged compared to the Norway case, except for a higher standard deviation for the case fatality rate and the hospitalization rate for severe cases. These numbers and their standard deviations are reported in Table 10.

**Table 10.**
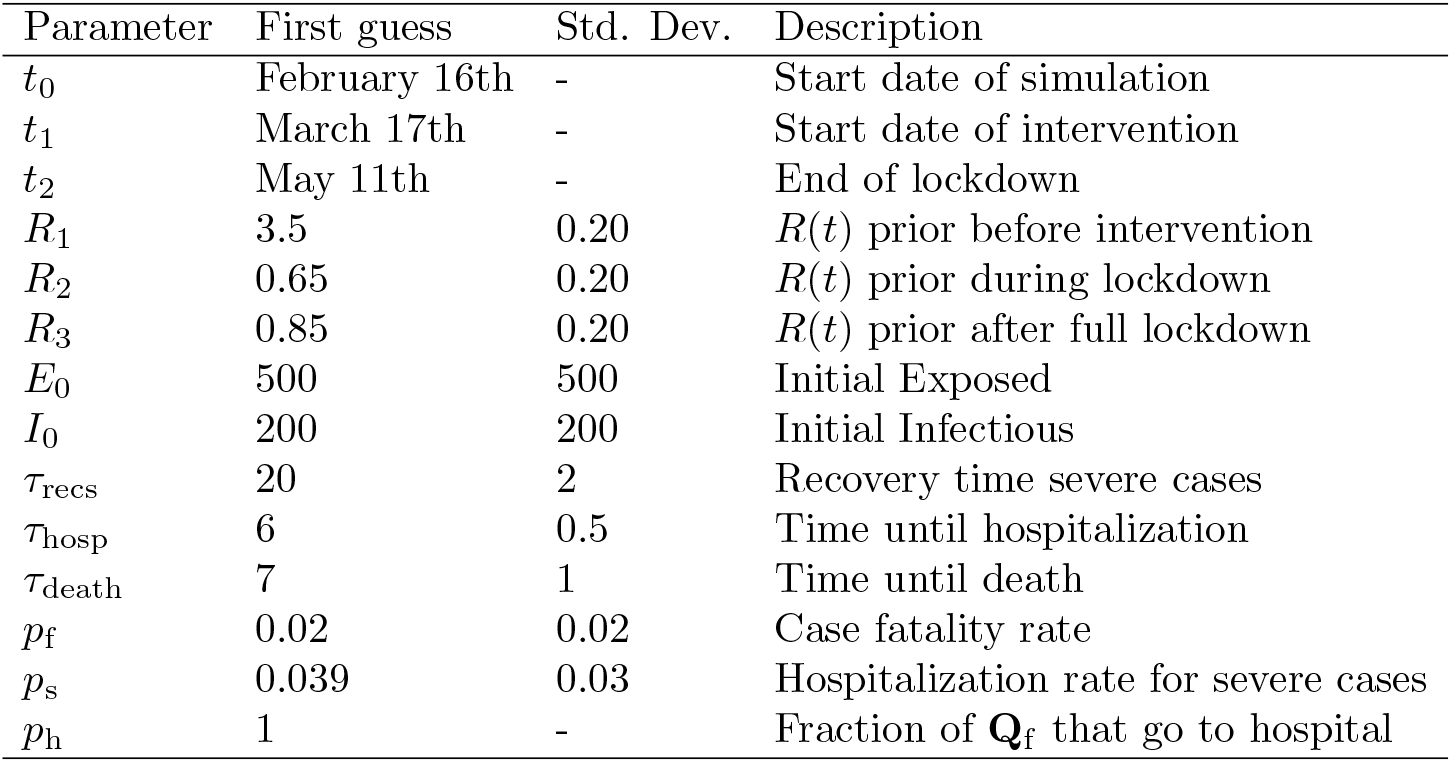
France: The table gives a set of “calibrated” first-guess (i.e., prior) model parameters and their standard deviations used for France. *p*_h_ is set ot 1 to inform the model that care homes deaths are excluded from the death numbers. All other parameter settings are unchanged as compared to the ones given in Table 1.

Figure 11 presents the assimilation results for the deaths at hospitals, hospitalized, and the reproduction number, as functions of time. The fit of our estimates to both sets of data is very good with thin posterior uncertainty. We have checked that such a performance cannot be achieved with the total death number (i.e., including the deaths in care homes). However, the estimated reproduction number before intervention struggles to remain close to the consensus value of about 2.5 to 3.5 [57].

**Figure 11.**
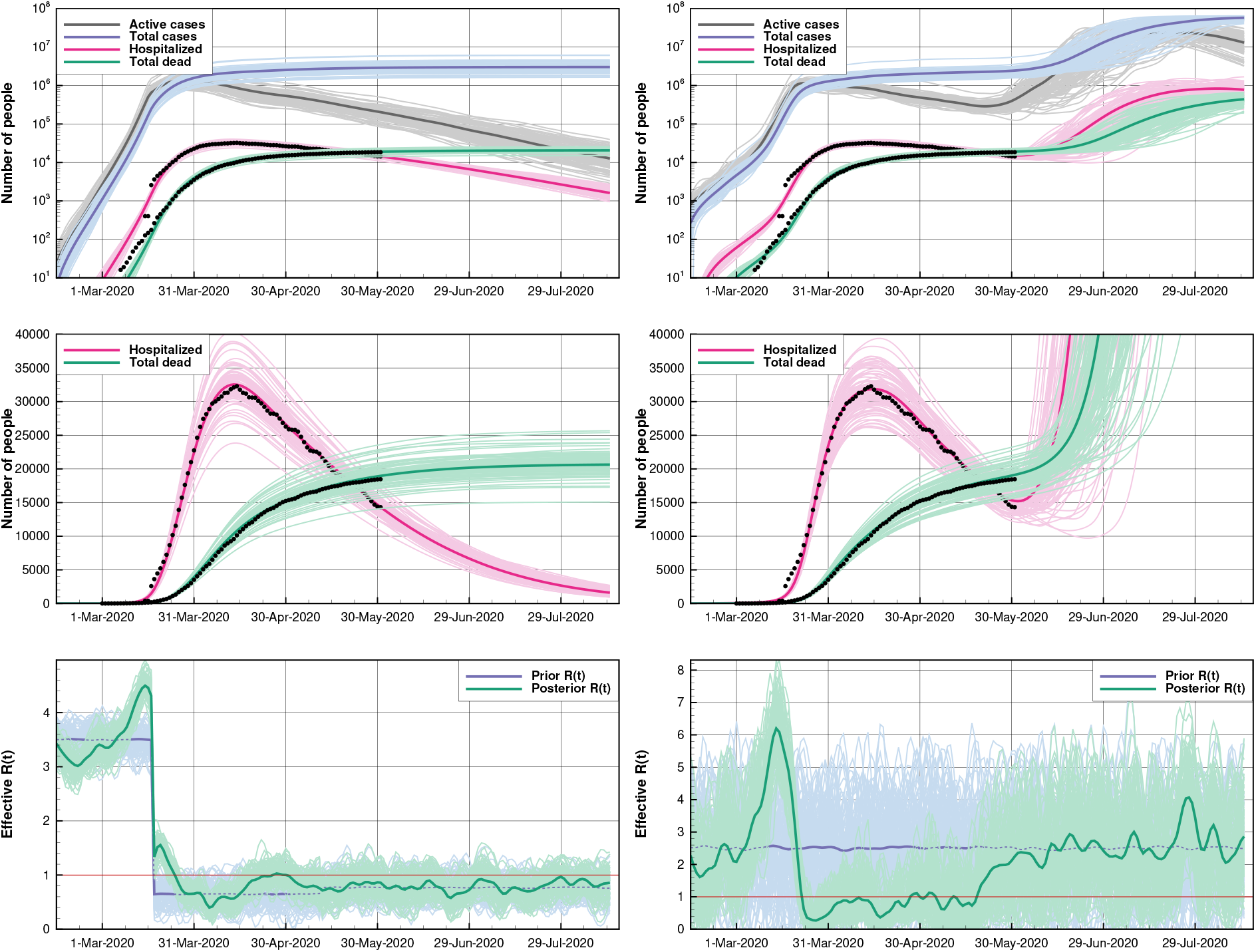
France: The figure shows the reference case (left) and case with an unknown intervention (right). The upper plots show the number of deaths at hospital, hospitalized patients, the total number of cases, and the currently active cases. The lower plots show the ensemble of effective reproduction number *R*(*t*).

The mean number of cases as of May 31st is estimated to be 3.66 ± 0.57 million people, which is about 5.47% ± 0.8% of the population. The fraction of the observed cases is estimated to be only 4.15% of the total number of cases.

### 8.2. Second experiment: no information on intervention

In this second experiment, we check if the data assimilation system can infer the lockdown period from the observations. The prior reproduction number *R*(*t*) is set constant in time at a fairly large first-guess value of 2.50 with a large standard deviation of 1.25, to make it as uninformative as possible. The other parameters are left unchanged, except for an adjustment of the initial Exposed and Infectious. The results for the time-dependent quantities are shown in Figure 11 (right panels). The fit to the observations, though not as good as in the first experiment, is reasonably good for both the accumulated deaths and the hospitalized. Furthermore, the algorithm detects a very significant change in reproduction number: *R*(*t*) between March 15th and 20th, and maintains it below one during the actual lockdown period. It shows that the lockdown was very effective in controlling the pandemic. The explosion in the death toll at the beginning of July (middle-right panel) is due to the high *R*_2_ prior and is not meant to be realistic.

### 8.3. Third experiment: forecast after lockdown

In this last experiment, we are interested in projections after the lockdown. The exit from lockdown happened on May 11th, but it was very progressive, with a second stage on June 2nd. The evolution of the situation after this date mainly depends on the scenario for the reproduction number, *R*(*t*). We have investigated three scenarios:

1. In the first scenario, the reproduction number is kept at a low value of *R*_3_ = 0.75. It is barely above the estimated value at the end of the lockdown, and it is compatible with the French government’s preliminary estimate on May 28th.
2. In the second scenario, the reproduction number is raised to *R*_3_ = 0.85, a somewhat ambitious targeted value.
3. In the third scenario, the reproduction number is raised to *R*_3_ = 1, the threshold case.

For this experiment, the setup is the same as the one used in the previous experiments. The results are shown in Figure 12. The two stable cases (*R*_3_ = 0.75 and *R*_3_ = 0.85), where the virus spread remains under control, yield very similar results, with about 21,000 deaths at hospitals by the end of August. By the end of August, it is possible to estimate the total number of deaths using the empirical ratio of deaths at hospitals to the total deaths computed from the French data as of May 31st. This ratio is about 0.64, so we predict that the total number of deaths at the end of August to be 33, 000. Finally, the *R* = 1 scenario departs significantly from the stable scenarios with a significant resurgence in the death growth (25,000 deaths at hospitals by the end of August 2020). These scenarios plead for maintaining a discipline of social distancing for a few additional months.

**Figure 12.**
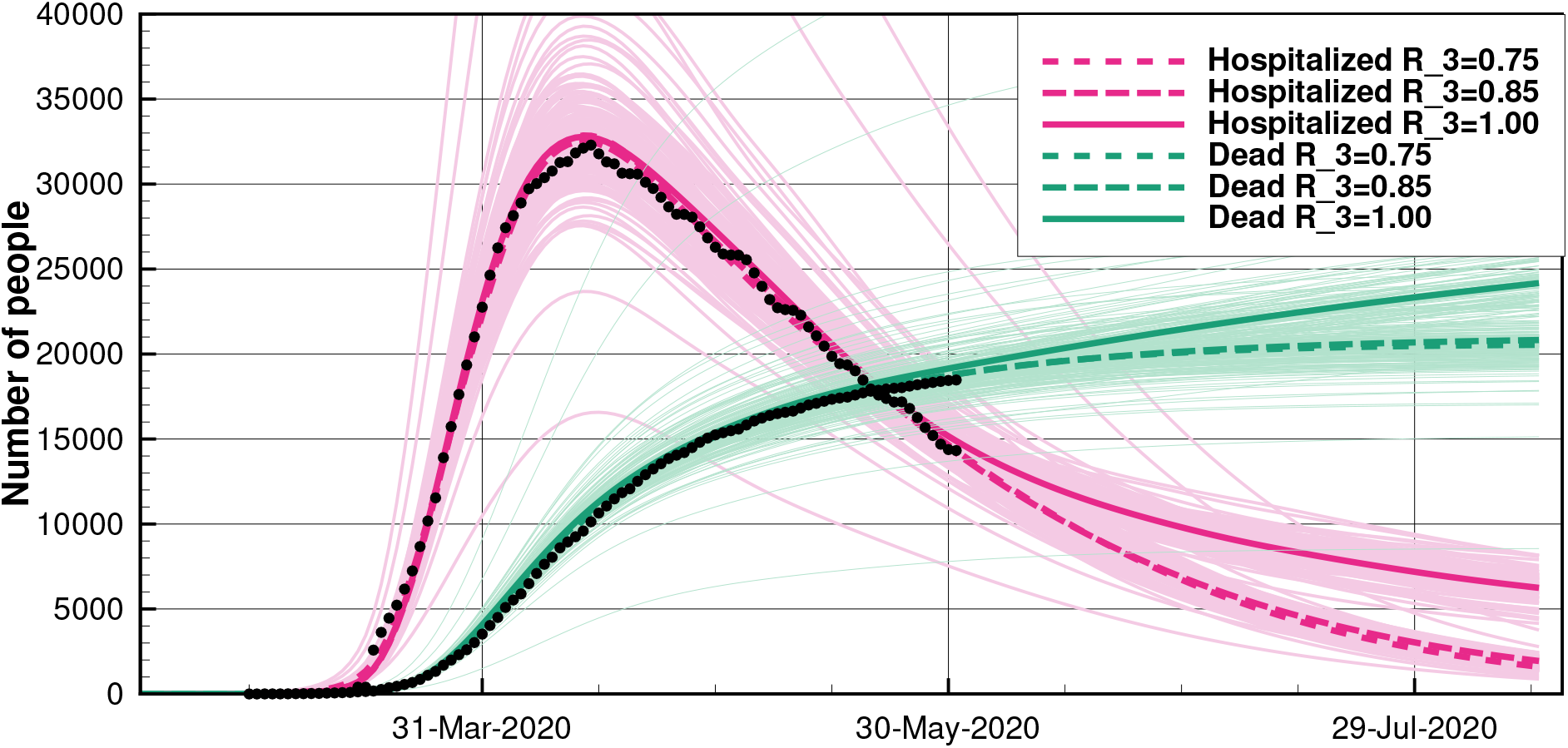
France: The plot presents forecasts of the reference case with three distinct scenarios after intervention setting the reproduction number to *R*_3_ = 0.75, 0.85, 1.00. We have plotted the posterior values for the number of deaths at hospitals (green lines), and the hospitalizations (red lines), together with the first hundred realizations for each case.

## 9. A case study for Brazil

We have employed the compartmental model, and the ensemble data assimilation described here to assess the possible future impacts on the relaxation of isolation policies and to illustrate the challenges involved in the prediction of COVID-19 spread in a vast country, mainly in the scenario of severe under-notification.

Many different sources (National Government, States Governors, and Cities Mayors) provide policy guidelines in a diverse country like Brazil. Therefore, it is a challenge to estimate the efficiency of, for instance, the distancing measures because of the different preventive policies adopted in each State [37]. For illustration, we have selected five different Brazilian states, one from each geopolitical region. In Figure 13, we show the evolution of COVID-19 in terms of number confirmed cases, deaths, and the mortality rate (CFR). The number of cases and deaths is still increasing in all the states, although at different growth rates and CFRs. For instance, there is a noticeable slowdown of the increase in confirmed cases in the Rio Grande do Sul (green curve). But not in Par (purple curve), where the confirmed cases have increased at approximately the same rate since the first death. It is also remarkable that the CFR tends to grow in time in Pará (purple curve) while it tends to decrease in time in Goiás (red curve).

**Figure 13.**
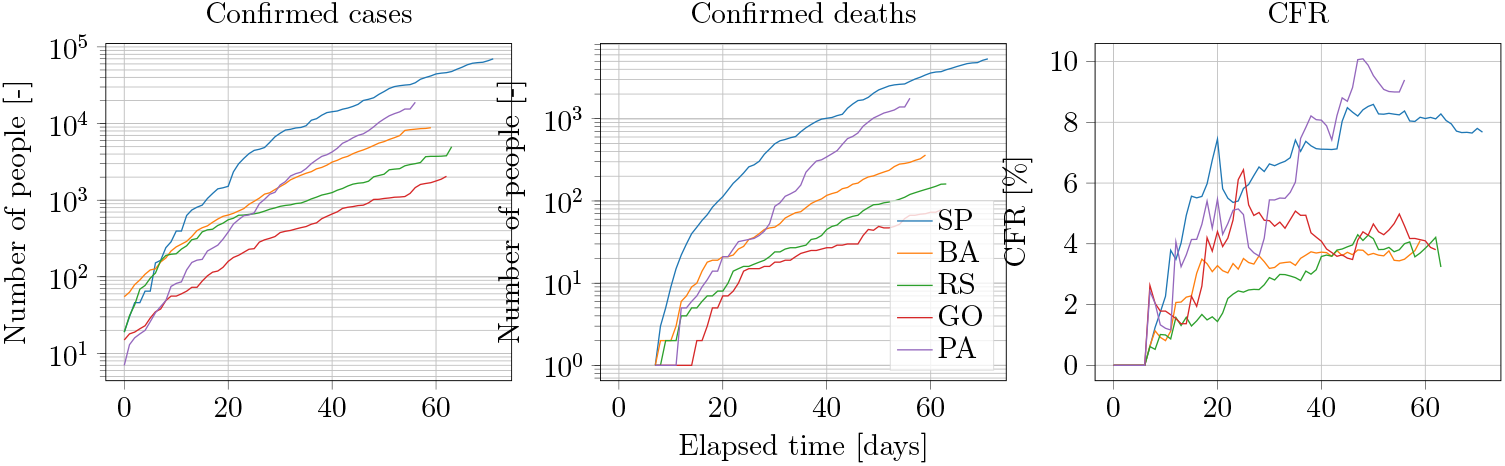
Brazil: The plots illustrate the evolution of COVID-19 in terms of the number of confirmed cases, deaths, and mortality rate (CFR), starting from the day of the first reported death by COVID-19. The reported states are São Paulo (SP), from the Southeast geopolitical region; Bahia (BA), from the Northeast; Para (PA), North; Rio Grande do Sul (RS), South; and Goiás (GO), Mid West. Each state represents a different evolution of the disease. We have shifted the curves in time to correspond to the first confirmed case.

Furthermore, there is a severe under-notification of both confirmed cases and deaths. The former is due to the lack of sufficient tests. The latter is related to a potential association of deaths, which might have been caused by COVID-19, to other respiratory diseases. Even though it has been reported an increased number of hospitalizations due to respiratory infections, substantially more substantial than other years, it is unclear what the actual number of admissions is due to COVID-19 [52].

For these reasons, we have modeled only one Brazilian State in this work, the State of São Paulo. This State concentrates the most significant number of confirmed cases and deaths in Brazil and still presents an increasing trend in the evolution of the disease. We focus on the spread of COVID-19 under the currently adequate quarantine level and in forecasting how the situation could be improved if more restrictive policies were applied.

The first decease due to COVID-19 in Brazil happened on March 20, 2020, in the State of São Paulo. Immediately after, on March 21, 2020, the State Government declared state-wide isolation. All commerce and non-essential services were to close on March 24, 2020. The intervention included all age classes, without any distinction, from closing kindergartens to any other establishment or social event that could result in large gatherings. The isolation policies were mostly followed and were still in effect when we ran the experiments.

For our data assimilation experiments, we make the following assumptions:

- Even after adopting a specific policy, its efficiency is uncertain. We assume that the policy results in a lower reproductive number, *R*(*t*), but it can still be higher than one (meaning the disease will still spread). As of mid-June, the epidemic is still growing, as is supported by a recent study [33, 39].
- The observation error on the number of deaths is primarily due to the under-notification. Thus, we will consider observation errors up to 20%, although they might be even more substantial.
- Due to under-notification, we will not assimilate the number of confirmed cases, where there is an even higher uncertainty around the reports [12].
- There are no observations of the number of hospitalizations due to the lack of reliable sources.

Table 11 presents the parameters used for the São Paulo State simulation. The reduced percentage of the hospitalization rate is due to the limited ICU capacity and due to the high number of deaths associated with severe respiratory diseases happening outside hospitals.

**Table 11.** Brazil: The table gives the parameter values used in the COVID-19 simulations for São Paulo, Brazil. All other parameters were kept unchanged as compared to the ones given in Table 1.

| Parameter | Initial value | Std dev |  |
| --- | --- | --- | --- |
| $t_0$ | March 10th | - | Start date of simulation |
| $t_1$ | March 23rd | - | Start date of interventions |
| $t_2$ | June 1st | - | Start date of prediction |
| $E_0$ | 656.0 | 65.6 | Initial Exposed |
| $I_0$ | 164.0 | 16.4 | Initial Infectious |
| $R_1$ | 4.0 | 0.4 | Prior $R(t)$ during spinup |
| $R_2$ | 1.0 | 0.1 | Prior $R(t)$ during interventions |
| $R_3$ | 1.0 | 0.1 | Prior $R(t)$ in prediction phase |
| $p_h$ | 0.2 | - | Ratio of fatally sick hospitalised |
| $p_f$ | 0.065 | 0.0065 | Case fatality rate (CFR) |

**Table 12.** US: The parameters used in our experiments, the different values for *R* at intervention steps are described in the sections for each case. Values without state indications are the same for all states. Any parameters not listed are the same as in Tab 1.

| Parameter | Prior | Std. Dev. | Description |
| --- | --- | --- | --- |
| $T_s$ | 20/2-2020 | - | Start Date |
| $E_0$ | 50(NY,CA),10(AL),20(NC) | 10(NY,CA,NC),2.0(AL) | Initial Exposed |
| $I_0$ | 10(NY,CA),1(AL), 2(NC) | 5(NY,CA,NC),1.0(AL) | Initial infectious |
| $p_f$ | 0.18(NY), 0.009(CA,NC,AL) | 0.001 | Case fatality rate |

Figure 14 illustrates the results of this scenario in the left plots. Since we allow *R*(*t*) to be higher than one, the number of deaths continues to grow until the end of 2020. The reproductive number before the first intervention is around four, consistent with the numbers reported by WHO. In the right plots of Figure 14, we show a similar data assimilation experiment, in which the reproduction number is kept at a low value of 0.6 after June 1st (simulating more efficient distancing measures). This measure could result in the ending of COVID-19 deaths before the end of 2020. The results illustrated in the left plots of Figure 14, show that, despite the isolation measures, we were not capable of reducing *R*(*t*) below one. Thus, the mitigation measures were probably not as restrictive as, e.g., in Europe. However, this situation can change in the future with the implementation of stricter rules. Thus, and it might be possible to reach a situation similar to the one illustrated in the right plots Figure 14.

**Figure 14.**
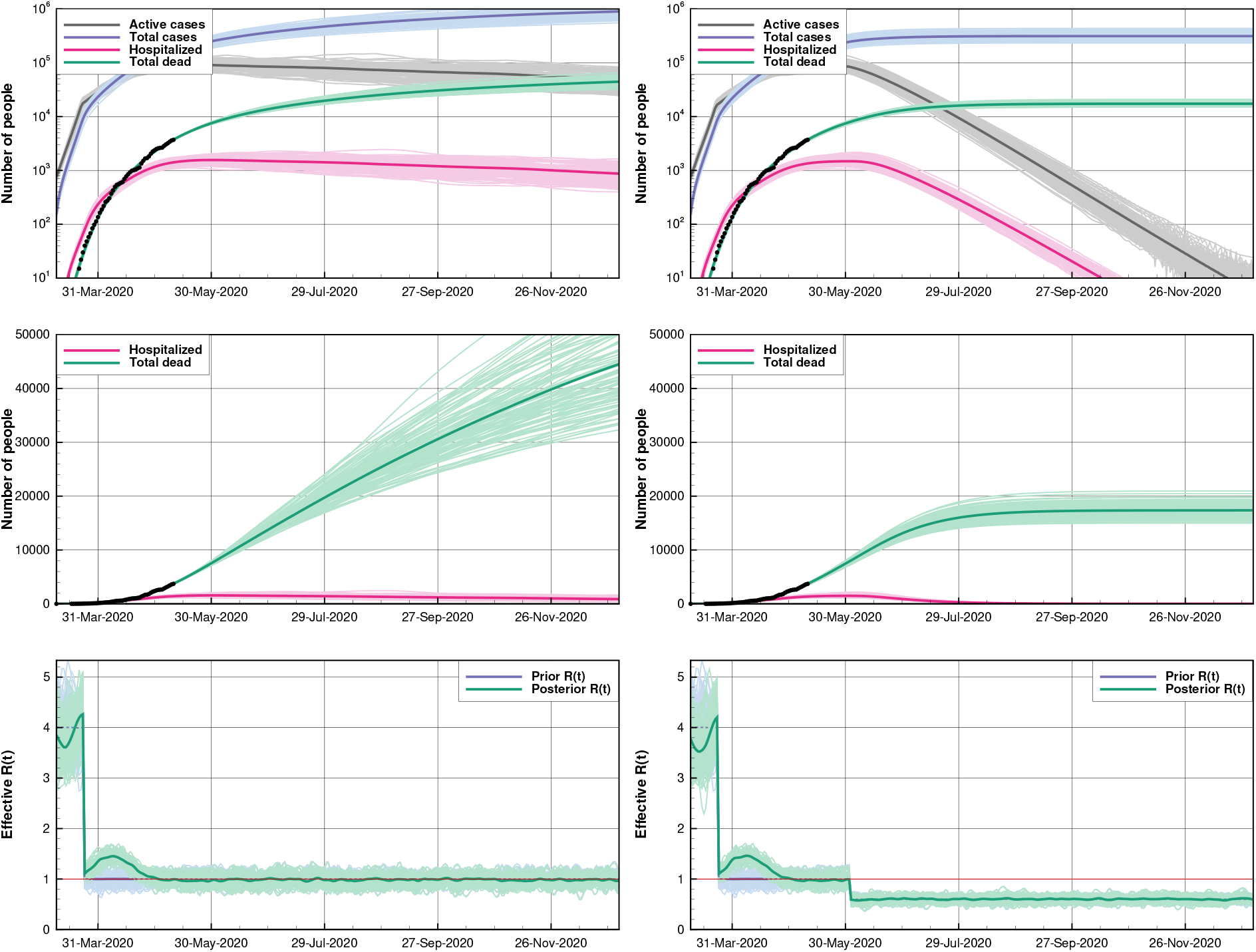
Brazil: The figure shows the simulation of the COVID-19 evolution in the São Paulo Brazilian State. In the left plots, we show a neutral case where the reproductive number *R*(*t*) ∼ 1.0 with a standard deviation of 0.1 for the prediction. The right plots present a stable situation where *R*(*t*) ∼ 0.6 with a standard deviation of 0.06, after a second intervention.

Next, we assess how uncertainty related to the effectiveness of interventions and under-reporting can impair policy-making. By setting different values and standard deviations on *R*_3_ (the prior for the prediction), we can simulate the impact of intervention uncertainty. The errors due to under-reporting can be examined by evaluating the prediction uncertainty due to observation errors.

We repeat the first and second experiments, but changing the observation error from 5% to 20%. The first and second rows of Figure 15 illustrate the results. Note that 20% errors can even be considered low according to some investigations that indicate a possible association of deceases reported to be due to other severe respiratory diseases to COVID-19 [52]. Also, we apply a standard deviation of 0.2 after June 1st for the neutral case, shown in the third row of Figure 15. This example illustrates the uncertainty around the actual measures assuming no changes in the current policies.

**Figure 15.**
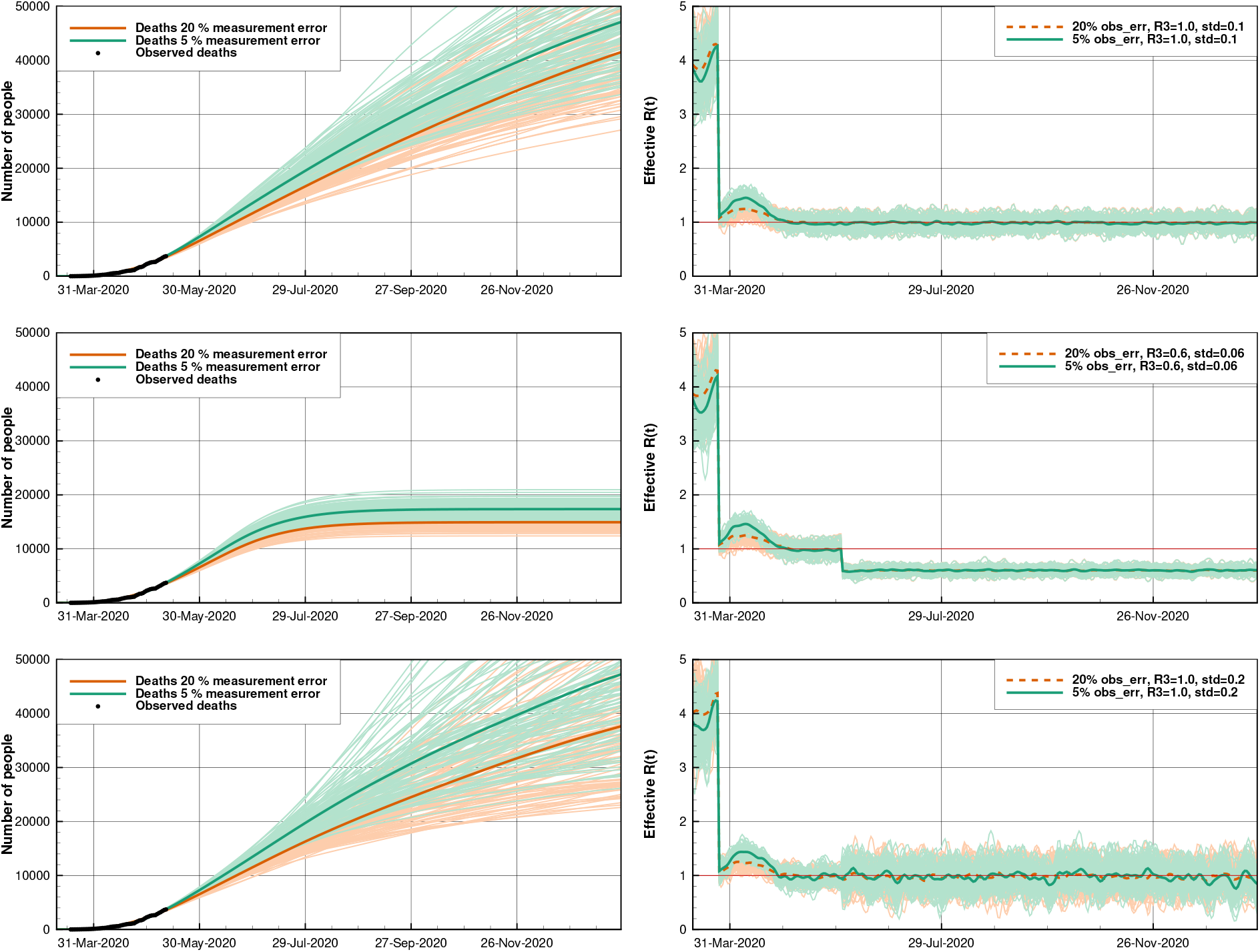
Brazil: The plots compare simulations considering relative observation errors of 5% and 20%. The upper and lower rows represent a neutral scenario with *R*_3_ = 1.0. They differ in the standard deviations or *R*(*t*) (0.1 and 0.2). The middle plots show a stable situation with *R*_3_ = 0.6 and a standard deviation of 0.06, following a second assumed intervention. The left plots show the deaths, and to the right, we present *R*(*t*) using different measurement errors.

It is incredibly challenging to construct, for instance, a plan to loosen up the isolation in the mid-long term, if the uncertainty around *R*(*t*) is high. This conclusion is supported by The broader spread of the posterior ensemble in the neutral scenario (*R*(*t*) ∼ 1.0) when a 0.2 standard deviation on *R*(*t*) is assumed and compared with the situation with a standard deviation of 0.1 (respectively, first and third columns in Figure 15). The wide range of plausible predictions represents a higher risk in any decision-making process. Note also that the stable scenario results in a narrower spread of the posterior ensemble (center plots). While this can lead to less risk, the more substantial observation errors still imply a difference of thousands of deaths, as shown in the expected mean curves in the center panel.

The evolution of COVID-19 can vary considerably depending on the geopolitical region or State. Additionally, even studies on individual states may not be entirely representative if we consider separate cities. Nevertheless, the significant uncertainty in the predictions, as in any other data assimilation or uncertainty quantification study, should be remedied by the acquisition of additional and more accurate data. More tests and a proper report of deaths, hospitalizations, and other vital data, are of utmost importance to reduce the uncertainty of the parameters, mainly on the reproduction number *R*(*t*), and hence in the predictions.

The unstable cases represent likely scenarios in case the government does not enforce policies to control the spread of the disease. Together with the stable examples, the results illustrate how the data assimilation system can support decisions to mitigate catastrophic outcomes by raising awareness and providing a tool for decision support.

## 10. A case study for Argentina

The SARS-CoV-2 virus arrived in Argentina around late February/early March after several citizens returned from the summer holidays, mainly from Italy, Spain, and the United States. Argentina detected the first positive case on March 3rd. Considering the large spread in Europe, including the situation in Italy and Spain, a group of epidemiologists advised the President on the need for social-distancing measures to slow down the spread of the virus. On March 20th, an extensive early lockdown in the whole country was established by the national government, at a time when the number of tested positive cases was rather small, i.e., about 100. This lockdown included strict control of the external country boundaries and between states, closing of all social activities and commercial businesses except shopping for food, and accessing the health service. Individuals were not allowed to leave their houses except for buying food. After about 40 days, in late April, the government started to relax the lockdown, permitting more work activities every week.

During March, the COVID-19 tests were collected around the country but only evaluated at a single place, Instituto Malbran, in Buenos Aires city. Because of this, there was a significant delay in the detection of cases of about five days. Furthermore, they were somewhat limited at first but increased later, which may change the fraction of undocumented cases and therefore affect the statistics. In late April, several authorized laboratories spread around the country started to analyze the tests. The Health Ministry produces a daily official report with the number of positive cases and the number of accumulated deaths. These reports are available on the Health Ministry web page. We used these data to conduct the data assimilation experiments of the virus transmission in Argentina. There is still some capacity available in the health system up to the end of May (Health Ministry web page), so that we consider the number of deaths due to COVID-19 to be rather precise. The standard deviation of the observed deaths was taken to be four at the start of the pandemic and increased up to 6 the last examined day, which contains an accumulated number of deaths of 539. Regrettably, the information on hospitalized people is not well updated on the available official databases, therefore we have been unable to use this information in the data assimilation experiments.

Since the first detected cases were around early March and because of the early lockdown, we conducted the data assimilation experiments for Argentina with a smaller number of initial exposed and infected individuals compared to the other countries, 40 and 10 respectively, and standard deviation of 8 and 2. We kept the rest of the parameters to be the same as for the other experiments (Table 1). We set the prior values for the effective reproductive number, *R*_1_ = 3.7, between March 1st and March 20th, *R*_2_ = 1.0 during the lockdown between March 21st and April 27th, and *R*_3_ = 1.5 after the relaxation of the lockdown.

We conducted two experiments, Case 1, where we conditioned both on the accumulated deaths and the reported number of cases, and Case 2, where we only conditioned on the deaths observations. Figure 16 presents the results from Case 1 in the left plots and Case 2 in the right plots, respectively. In the upper plots, we show the reported number of accumulated cases and deaths as dots. For the number of cases, we assumed that this number represents only 15% of the actual number of cases. Since there is no observational evidence in Argentina about the reported total cases rate, we assumed the estimate from Chinese data by [38]. The thick lines correspond to the posterior density mean of the total and active cases, the number of hospitalized individuals, and the number of accumulated deaths. The estimated mean parameters exhibit only slight deviations from the proposed prior values, except for the case fatality rate, which we compute to be *p*_*f*_ = 0.011.

**Figure 16.**
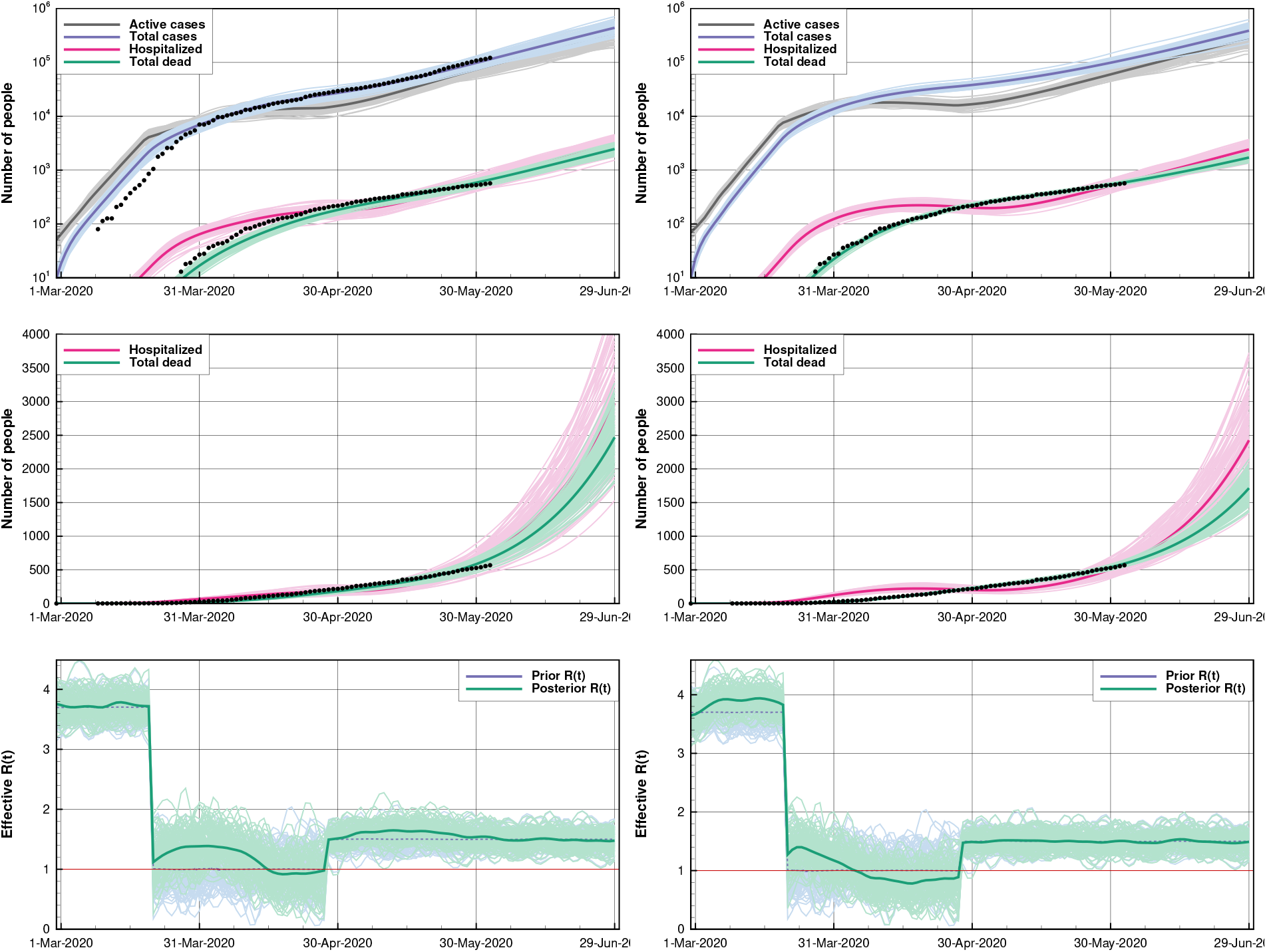
Argentina: The dots are the observations. In Case 1 (left plots) both the accumulated deaths and the estimated number of cases were conditioned on, while in Case 2 (right plots) we only conditioned on the total number of deaths.

In the lower plots of Figure 16, we present the mean (line) and uncertainty, represented by the ensemble, of the effective reproductive number *R*(*t*) as inferred with ESMDA. Overall, the estimated *R*(*t*) in the pre-lockdown period is 3.7 and rather close to the assumed prior mean. After the lockdown, *R*(*t*) descends abruptly to about 1.1. After that, we see a slight increase in *R*(*t*) to over 1.5 following the relaxation of the lockdown. Note that the estimated *R*(*t*) in Case 1, corresponds to a minimal number of reported cases and deaths. The early lockdown effectively diminished the increase in the number of new infections and the number of hospitalized and fatalities. Two months after the first reported COVID-19 case, the number of reported cases was still relatively small, 4783.

The early lockdown did reduce *R*(*t*) below one for a short period, but the relaxation of the quarantine led to values over one. Thus, the number of cases is still increasing as of the end of May, and the outbreak’s peak is ahead. We partly attribute the lower reduction of *R*(*t*) compared to other countries to the social inequality distribution in South America. The lockdown was not effective in vulnerable neighborhoods because of the living conditions. In those disadvantaged neighborhoods, many individuals are living in the same small house, and most of the daily activities are conducted outside the house so that strict social distancing measures are not viable. The number of persons detected with COVID-19 in these neighborhoods has increased dramatically with time. Meta-population or agent-based models focusing on the neighborhood scale are required to model these patterns, in particular the spread’s spatial distribution.

Since the reported number of cases by the health system may be underestimated, we conducted an experiment (Case 2) in which we use only the number of deaths as observations. In this case, we obtain a closer match to the observed number of deaths. On the other hand, the inferred number of reported cases is larger than the reported cases. This result may indicate that the number of tested cases was lower than 15% of the total cases. Still, it may also be the result of the delay in the evaluation of the tests. As shown in the middle plots of Figure 16, the posterior mean of the accumulated deaths shows a closer match to the data. The effective reproductive number resulting from conditioning only on the number of accumulated deaths shows significant differences to the results from Case 1. The estimated mean exhibits a lower level of variability. It descends to values below one during the lockdown period, and then it increases to 1.4-1.5 after the relaxation of the lockdown. This behavior produces a flattening of the hospitalized number compared to Case 1.

Because of the long time lag of death number compared to the reported cases, the estimated *R*(*t*) using the reported cases has more recent information about the infections. Indeed, Case 1 captures a change in the trend of the last infections with an increase in *R*(*t*) to values of 1.6-1.7, which is still not detected in Case 2 (Figure 16). In this sense, reported cases may have some valuable information for producing forecasts. However, the correct trade-off between different kinds of observations must be found since the reported case numbers are less reliable than death numbers.

To evaluate the system as a tool for decision making, we conducted a third experiment focused on probabilistic forecasts. As in Cases 1 and 2, we started with the prior of *R*(*t*) = 3.7 until the introduction of interventions on March 20th, where the prior for *R*(*t*) is reduced to *R*(*t*) = 1.3 until June 2nd. We then modeled three potential scenarios with *R*(*t*) = 1.7, 1.3, and 0.9, representing a further relaxation of the lockdown, a continuation of the current partial lockdown and social distancing recommendations, and a more restrictive scenario with a return to the original lockdown measures. Because these are probabilistic scenarios, the standard deviation of R(t) was reduced to 0.05. The upper plot of Figure 17 shows the evolution of the number of deaths and reported cases while the lower plot presents the effective reproductive number. In these scenarios, we only condition on the numbers of dead, which are more reliable than the cases. The probabilistic predictions differ substantially in the three scenarios. On August 1st, the predicted mean number of deaths is 6800, 2800, and 1600, respectively, while the forecasted estimates of reported cases are 302000, 73000, and 27000 for the three scenarios. These are only indicative values. A regional study would probably provide for a more precise long-range prediction of the pandemic evolution.

**Figure 17.**
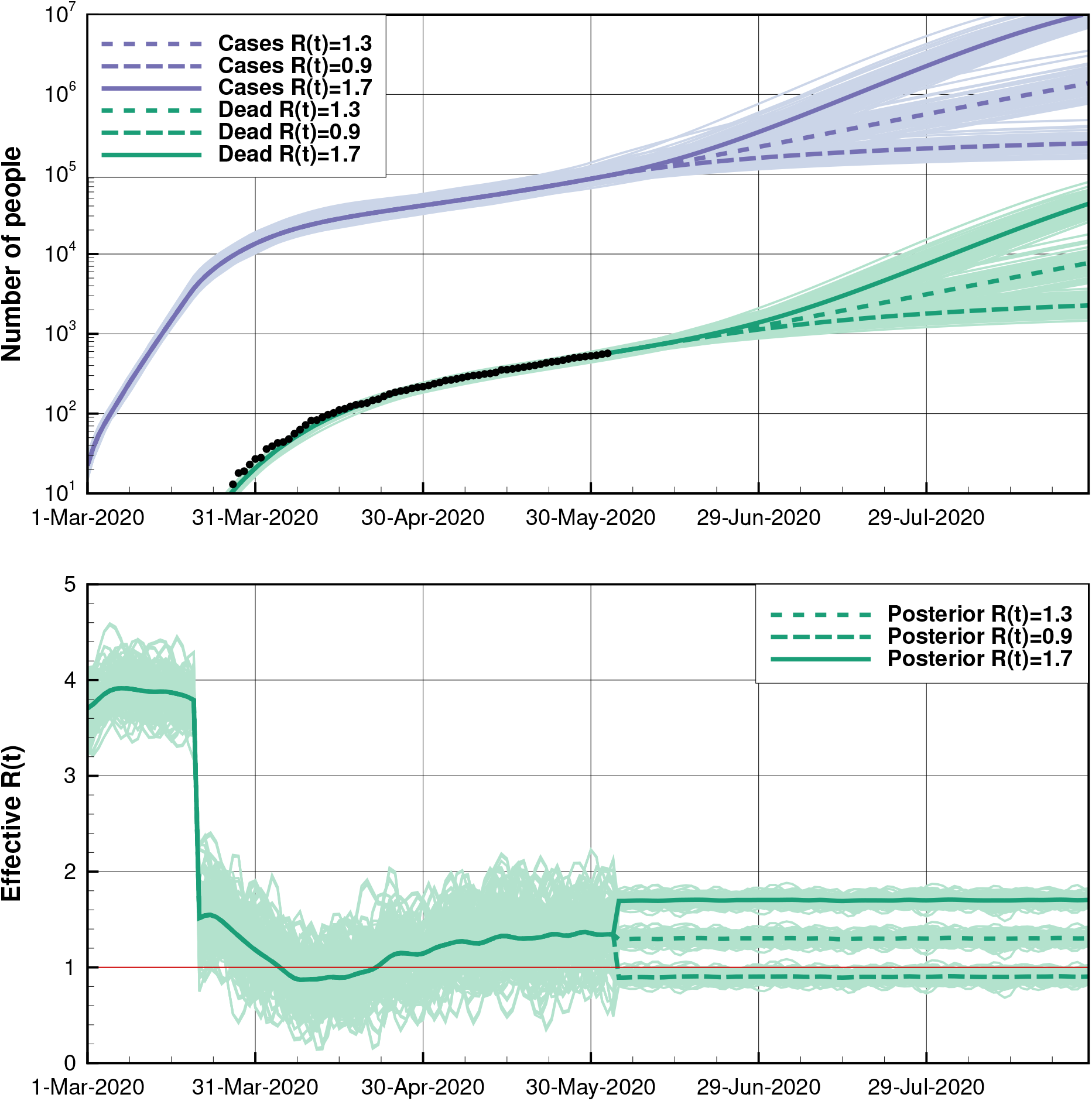
Argentina: Same as Figure 16 but for an experiment focused on a probabilistic prediction in which three scenarios, with different effective reproductive numbers imposed from June 1st, *R*(*t*) = 1.7, 1.3, and 0.9.

## 11. A case study for The US

The novel SARS-CoV-2 virus was in the early stages of community spread in late February when the first non-travel related case was discovered on February 26th in California. An analysis of RNA sequences from early cases suggested that a single lineage of the virus was imported directly or indirectly from China, followed by several importations of SARS-CoV-2 from Europe [36]. The largest number of flights between The United States and Europe typically go through airports in the northeast region of the country, particularly in and near New York City. A lack of testing led to some uncontrolled spread early on as individual states began their first mitigation measures.

These initial measures, such as school closures and gathering size restrictions, were taken around March 10th for most of the states. We can see an impact of these measures in Figure 18 by the initial drop in mobility, measured from cell phone data and compiled by the Institute for Health Metrics and Evaluation (IHME) [41]. On March 16th, the Center for Disease Control (CDC) and the newly formed White House Corona Virus Task Force announced voluntary guidelines dubbed “15 days to slow the spread.” These guidelines were later extended through April. They advised taking only essential trips combined with some social distancing measures but, with a focus on following specific instructions of the individual state governments. As a result, each state took its path with varying degrees of restrictions and their enforcement. The response of the people of each state varied as well. Protests against stay-at-home orders sprang up in many states, closed beaches were sometimes filled, and some churches defied closure orders.

**Figure 18.**
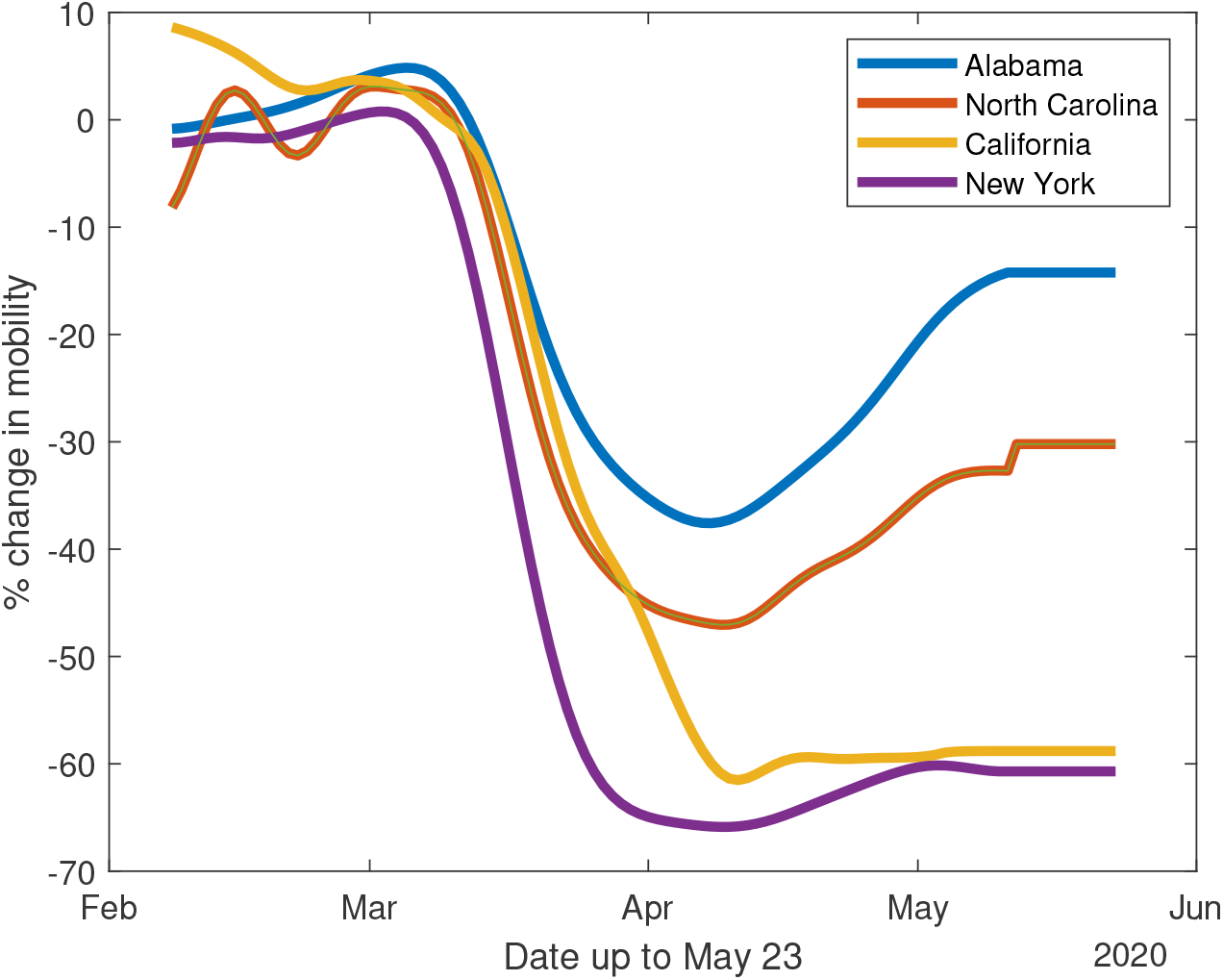
US: Mobilites for each of the states considered

With this backdrop of heterogeneity across the US, we carried out a case study of four states representing a cross-section of the country: New York, California, Alabama, and North Carolina. New York was chosen for two reasons. First, it was the hardest-hit state due to the population density in New York City as well as its strong connection to Europe through air travel. Secondly, based on mobility data, New York achieved a 66% reduction from baseline mobility, one of the most substantial decreases observed in the United States. California was chosen for similar reasons, with an achievement of a 55% reduction in mobility. It was also one of the quickest states to have, at least parts of it, lock down. On the other hand, it also experienced massive protests and defiance of beach closures. Alabama and North Carolina are chosen because of lower reductions of baseline mobility of 38% and 47%, respectively, and their contrasting approaches to government mandated measures despite similar demographics.

We assimilated the number of accumulated deaths and current hospitalizations using three different scenarios. First, we set the prior for the reproduction number to be *R*(*t*) = 1 with a large uncertainty to see how the ESMDA estimates *R*(*t*). In all of the cases, the prior of *R*(*t*) = 1 does not appear to be viable until around late March, which is consistent with the mobility data approaching their respective minimums around that time for all four states.

Secondly, we assume that every citizen followed the CDC’s guidance issued officially on March 16th, causing an immediate drop in the reproduction number to *R*(*t*) = 1. And thirdly, we implemented a step-down decrease from a maximum *R*(*t*) value to an intermediate value and then to *R*(*t*) = 1. We choose the date for setting the prior of *R*(*t*) to its intermediate value, to be the first date when the mobility data reaches half of its maximum reduction-value, typically around March 17th. After that, it follows a transition to *R*(*t*) = 1 when the mobility is ten percentage points away from the maximum reduction value, typically around March 23rd.

We have chosen the prior *R* values at the first two steps so that the forecast ensemble mean closely follows the number of hospitalizations and deaths. We set the final prior of *R*(*t*) to one so that we can gauge the success of the mitigation efforts for the given state. The time-varying value for *R*(*t*) is updated in the assimilation step and implies that when *R*(*t*) > 1, after the final step down, the state has not met its mitigation goals. Similarly, when *R*(*t*) < 1, the state may have exceeded its mitigation goals.

### 11.1. Data and Parameters

The data we use comes from the IHME group at the University of Washington. They have pre-compiled the historical data in a large CSV file, which we use to extract data for the 50 states: http://www.healthdata.org/covid/data-downloads. Their primary data source is John’s Hopkins University. We use the accumulated number of deaths, the current number of COVID hospitalizations, and the accumulated number of positive cases. However, we do not condition our ESMDA steps on the number of cases due to the lack of early testing in the United States as well as considerable uncertainty in the percentage of cases it represents. Our population data for each state comes from the 2018 U.S. Census Bureau estimates. We use an age grouping similar to the Norwegian case with 11 age groups.

### 11.2. Case 1: Sensitivity analysis on *R*(*t*)

In this case, we take a prior of *R*(*t*) = 1 and increase the uncertainty in *R*(*t*) to have a standard deviation of 1 to see how the data may inform the reproduction number with the results shown in Figure 19. In this case, we condition on deaths and hospitalizations and observe how *R*(*t*) differs from a base prior value of *R*(*t*) = 1 over time. Both New York and California show a sharp rise and then decrease with New York having the largest values for *R*(*t*) before the implementation of measures. Both states issued similar rules at similar times but have very different urban structure. People in New York City often use crowded public transit, while in Los Angeles and other cities in California, personal vehicles tend to be the primary mode of transportation. North Carolina has initially slower spread but is comparable to Alabama in the early days with larger oscillations around *R*(*t*) = 1 after late March than California or New York. While this analysis is informative to give us an idea of how the virus was spreading in time, we also want to condition on priors that represent specific scenarios *intended* by lockdown measures. We consider these types of priors in the next two cases.

**Figure 19.**
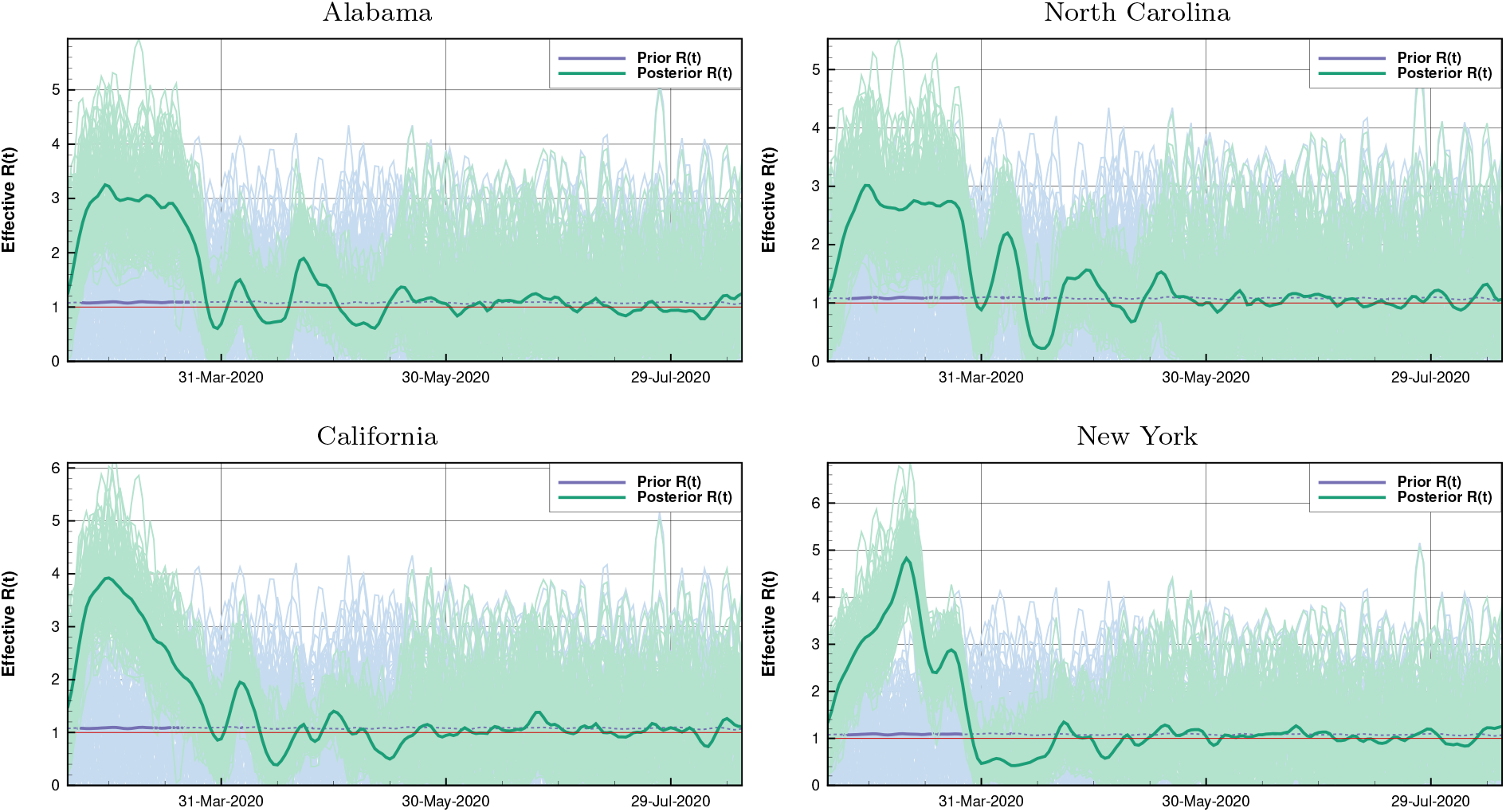
US Case 1: Large uncertainty in *R*(*t*)

### 11.3. Case 2: Immediate Response to Federal Guidelines

In this case, we impose a drop in the prior in *R*(*t*) to take place when the first federal guidelines were issued on March 16th with the results shown in Figure 20. Clearly, from the mobility data in Figure 18, this drop is unrealistic as mobility decreased slowly from mid-March to mid-April. However, it is essential to establish that the assimilation of deaths and hospitalizations detects this decrease, as, for all cases, *R*(*t*) remains well above one until the mobility indeed decreases enough. New York shows a sharp decrease in *R*(*t*) in late March while the other three states lag until the beginning of April. Interestingly, in North Carolina, the value for *R*(*t*) remains relatively high until around April 15th, peaking on March 31st despite that the mobility having decreased significantly during the weeks before. A reason might have been an influx of people from New York and the northeast with second homes in North Carolina. The sharp drop in mobility in the state of New York is also apparent in this analysis. What is clear is that New York had the most consistent and rapid response in terms of controlling the reproduction rate.

**Figure 20.**
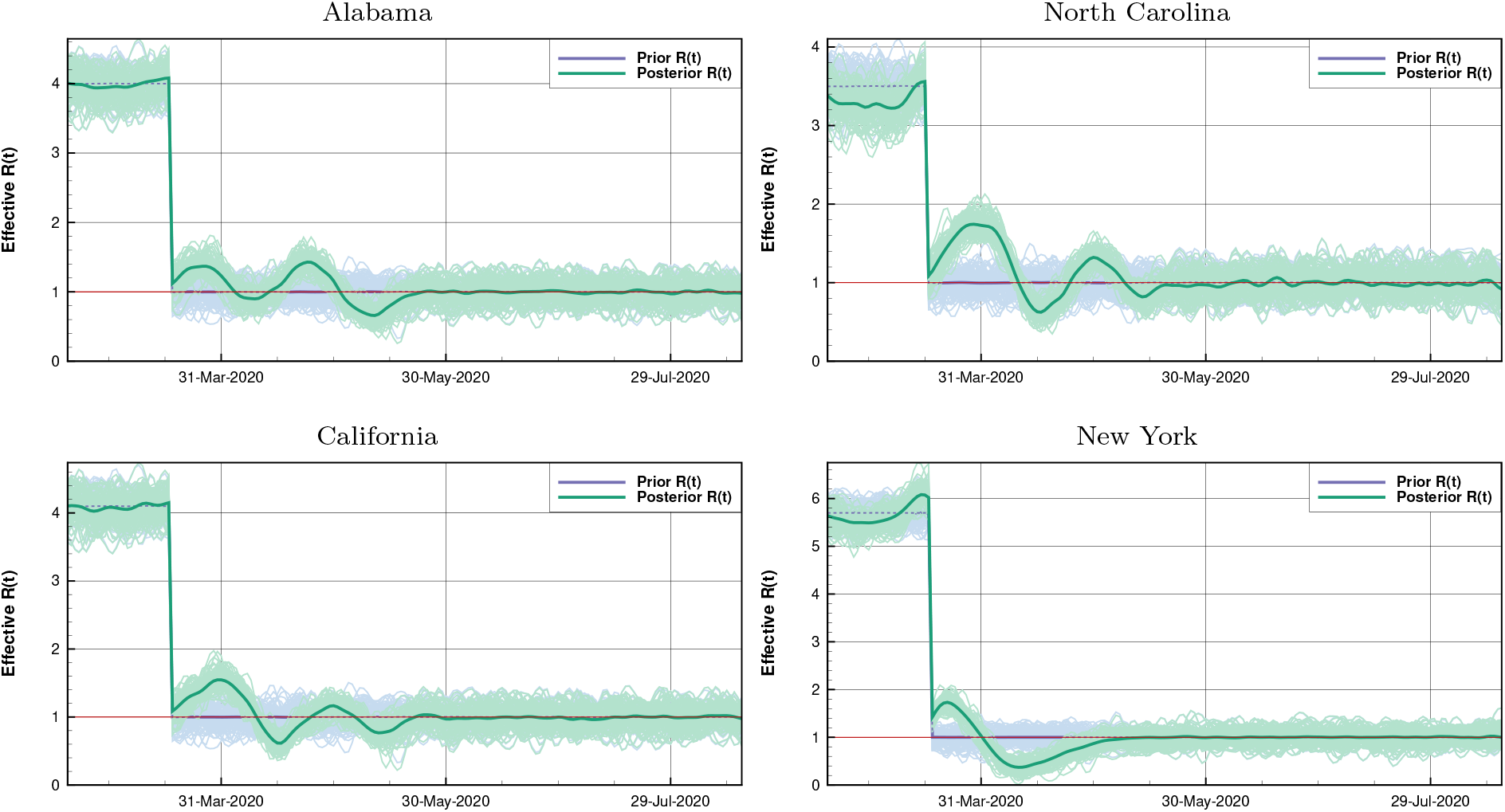
US Case 2: Assuming voluntary federal guidance was immediately observed by the citizens on March 16th.

### 11.4. Case 3: Intermediate step down to *R* = 1

In this case, we impose two drops in the value for *R* to better account for the slow decrease shown in the mobility data with the results shown in Figure 21. We choose the first two values for *R*(*t*) so that the initial forecast mean closely follows the data on deaths and hospitalizations until the step down to *R*(*t*) = 1. In this way, we can gauge how well a state has managed to control the spread of the virus, and compare the projection of the “goal spread,” (achieving *R* = 1), and the projection influenced by the data. It is immediately clear from the effective *R*(*T*) ensemble plots that neither Alabama, California or North Carolina were able to achieve constant control of spread. Alabama has some of the least reduction in mobility with the longest lag time in reaction to the federal guidance. It is notable that before the intervention, the *R* value for Alabama fit fairly well at *R*(*t*) = 4 while for North Carolina *R*(*t*) is lower at just below 3.5. This is despite the two states having a similar geographical distribution of the population. Further, Alabama typically had *R*(*t*) above one with a dip below one shortly after the minimum mobility. The difference in *R*(*t*) values between California and New York before intervention is also striking. New York fits well with values as high as *R*(*t*) = 5.7 while California hovers around *R*(*t*) = 4 in the early days. Both states have large Urban areas with large populations; however, the population density in the urban centers in New York, NYC, in particular, is typically much higher. There are differences in the methods of travel as well, public v.s. private transportation as discussed in Case 1. With all states but New York, we see oscillations around *R*(*t*) = 1, which may be linked to specific spreading events. California and North Carolina experienced a series of anti-lockdown protests. The largest protest in California, on May 1st, included more than 1000 people. It was one out of a series of protests numbering in the 100s, in the days before and after May 1st. In North Carolina, there were two protests, 100 people on April 14th and 300 people on April 21st. New York also experienced two protests, one low risk protest in Albany on April 22nd, where a parade of cars drove through Albany’s capitol park and one on May 1st where hundreds of protesters converged on Commack, Long Island. It is apparent, however, that New York achieved sustained *R*(*t*) below *R* = 1 and then stabilized around *R* = 1 as in the last half of May with the data going up to May 23rd. By and large, the citizens of New York were far more cooperative than in many other states, but this could also be an artifact of the sheer number of infections and deaths New York suffered in the early days of its outbreak.

**Figure 21.**
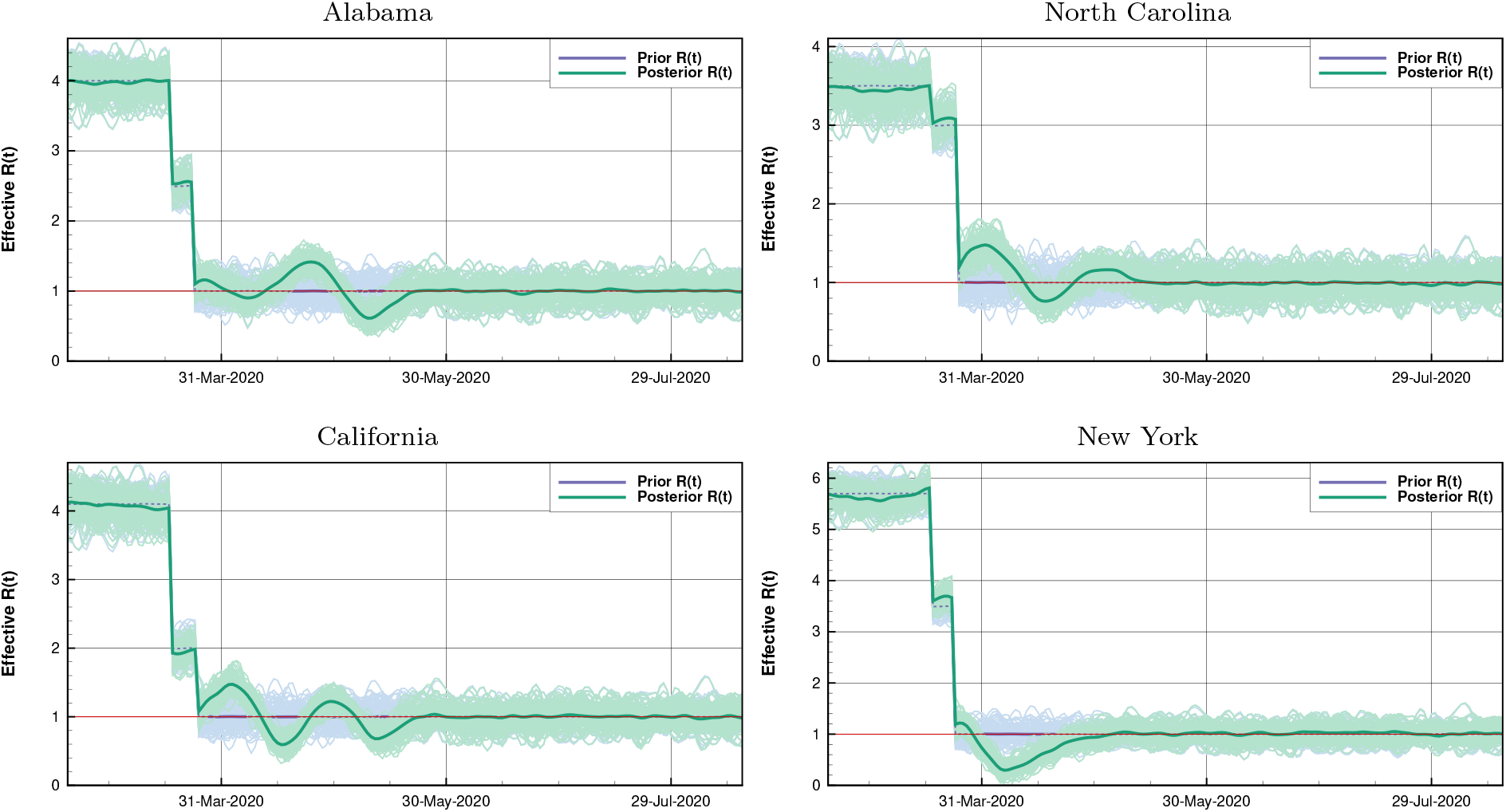
US Case 3: A gradual step down in *R*(*t*) with the first and intermediate values chosen so that the prior mean closely follows the data until the time for which *R*(*t*) is guessed to be one.

We consider the step-down runs to be the most realistic in terms of forecasting for each state. In Figure 22, we show both the mean of the forecast, chosen to closely match the data on hospitalization and deaths, as well as the analysis projections before transitioning to *R*(*t*) = 1 as our prior. The projections past May 23rd, the last day of available data, in this case, rely on *R*(*t*) = 1 for the rest of the year. It can be seen that California, North Carolina, and Alabama will not do better than the situation represented by the forecast mean in terms of accumulated deaths. However, New York is on track to doing better than the *R*(*t*) = 1 prognosis after the last intervention due to its sustained *R*(*t*) < 1 through mid-April. If the other three states manage to maintain some *R*(*t*) ≈ 1, the variations in the early days of the lockdowns do lead to more deaths than otherwise would have occurred. Furthermore, because *R*(*t*) remained above or around *R* = 1, no rapid decrease in cases was achieved. The time to reach zero hospitalizations has increased significantly and projects to continue through Summer 2020. On the other hand, New York is currently on track to end the outbreak by September.

**Figure 22.**
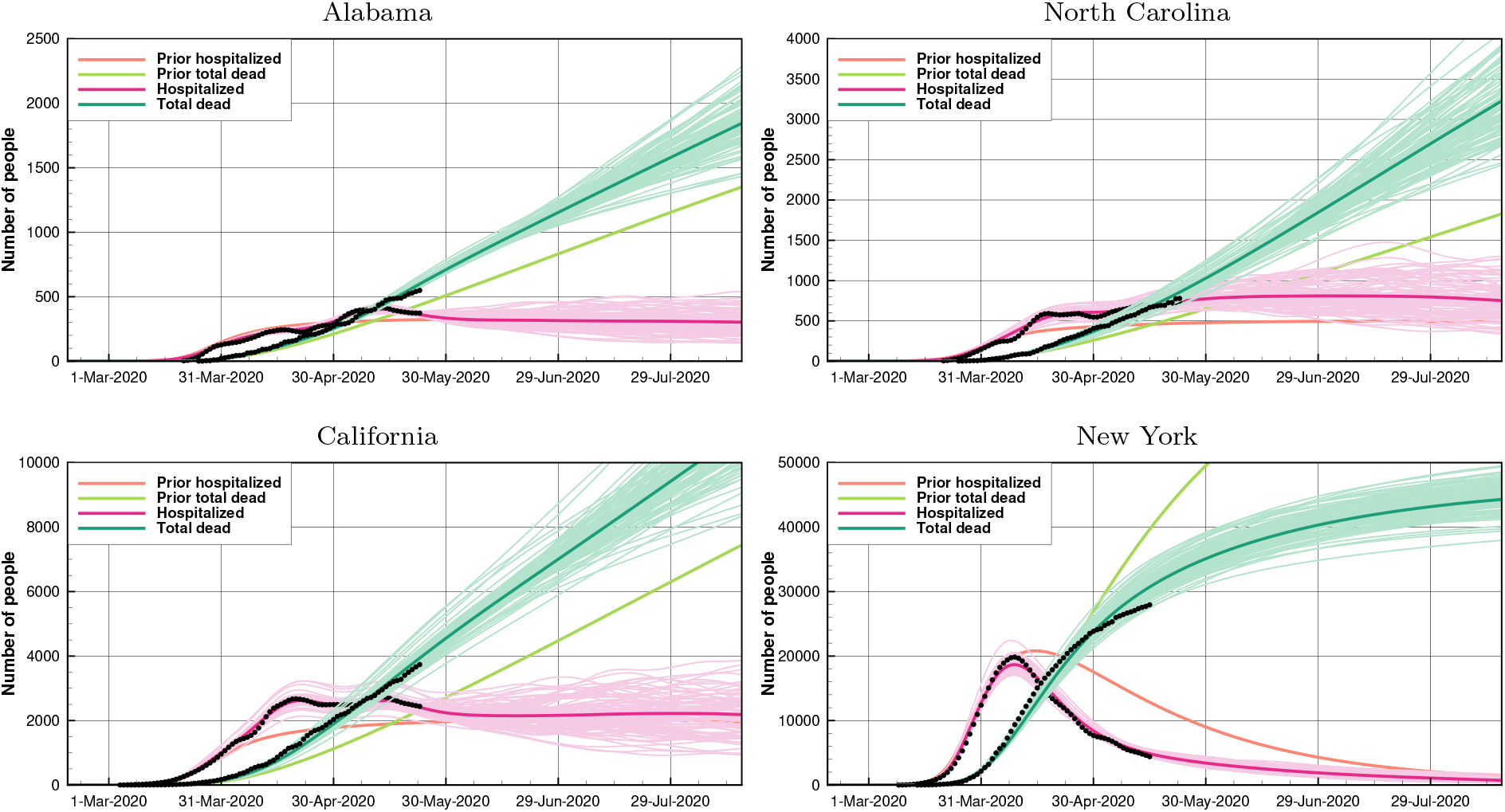
US: Forecasts for Case 3: Here we also show the prior forecast mean to highlight the difference between the idealized scenario and after analysis. In this case, only New York was able to achieve a trajectory that predicts fewer deaths than the idealized scenario. Furthermore, New York also is the only state in this study to make an end of the outbreak by late August.

## 12. Discussion

We have demonstrated an approach for parameter estimation, and model calibration in an age-stratified SEIR model used to model and predict the COVID-19 epidemics. By assigning uncertainties to all poorly known parameters, and characterizing them by specifying appropriate prior distributions, we can sample large ensembles of parameter realizations. The model predictions originating from the prior ensemble of parameters constitute our previous knowledge of the epidemic evolution, before data are taken into account, and have typically significant errors. The use of an ensemble data assimilation method allows for estimating both static model parameters and the time-varying effective reproductive number. Our ensemble data-assimilation approach, conditioning the model on observations, implies constraining the prior ensemble, and compute an updated ensemble of parameter estimates that constitute the posterior solution. The posterior solution agrees well with the measurements and has a significantly lower uncertainty. As such, the use of ensemble data assimilation allows for efficient model calibration, and we propose it as the ideal tool for calibrating any epidemic model to observations.

### 12.1. ESMDA convergence and ensemble size

It is possible to interpret the ESMDA method as a sequence of Euler steps in the parameter space, starting from the prior and ending up at the posterior solution. The technique applies a linearization at the beginning of each pseudo-time step to determine the update-direction. Thus, by reducing the step size, one would expect that the accuracy improves. Therefore, independent of the ensemble size, we need to use sufficiently short steps to ensure that we have converged to the solution of the method. Previous studies using ESMDA have indicated that the technique requires about 16–32 steps to converge with reasonable accuracy. Thus, we have used 32 MDA steps in all the experiments presented in this paper.

An ensemble size of 100 realizations will provide a reasonable solution, but there will be significant sampling errors. A sensitivity study indicated that using more than 5,000 ensemble members will not lead to visible changes in the results. Thus, we have used 5,000 realizations for all the final cases presented in the paper.

A document with supplementary plots, illustrating the convergence of ESMDA, is available from the Github repository https://github.com/geirev/EnKF_seir/doc/sensitivity.pdf.

### 12.2. Model initialization

Given a prior set of model parameters, e.g., the defaults in Table 1, we can initialize the model by setting the start date, the initial number of infectious *I*_0_, and exposed *E*_0_. These numbers interact with the reproduction number *R*(*t*) for the initial period between the start date and the introduction of the interventions. As the value of the basic reproductive number is itself somewhat uncertain, we have allowed for uncertainty in *R*(*t*) in the initial period. Given a start date, we have some freedom to balance *I*_0_, *E*_0_, and *R*(*t*) in the initial period, such that we match the observed onset and numbers of deaths and hospitalized. There are different combinations of these parameters that will give an equally good fit to the observations. However, the critical point is to have a realistic model estimate at the onset of interventions, and the initial period serves mostly as a spinup period. The data will, after that, constrain the system.

In several of the cases presented here, we used ESMDA to estimate these initial parameters in a recursive process to find a good initial model state. After that, we initialized the data assimilation experiments with these prior values using smaller uncertainties but added uncertainty to the additional model parameters.

### 12.3. Estimation of *R*(*t*)

A strength of the method is that it is also possible to estimate the effective reproductive number as a function of time. The cases in which we choose a constant prior *R*(*t*) (for The Netherlands, France, and The US) and a high standard deviation to reflect the uncertainty, the observations steer the posterior estimate of *R*(*t*) to more realistic values. In this manner, the data assimilation can be used in a hindcast mode to provide an analysis of *R*(*t*) and test the validity of different scenarios for its temporal evolution.

All experiments that have a sudden lowering in the prior value of *R*(*t*) exhibit a compensation effect shortly after this change. By implementing this reduction over two smaller steps rather than one, as in Case 3 for The US, this effect is less, but still present. Using a more gradual *R*(*t*) as in Case 3DH for The Netherlands results in a smoother *R*(*t*) but does not eliminate the tendency of the system to produce a low value of *R*(*t*) following a peak.

Since it takes about two weeks before new cases evolve into hospitalizations or deaths, it is possible to constrain or estimate *R*(*t*) until about two weeks before the latest observation. For the last two weeks and into the future, *R*(*t*) will stay close to its prior value. Consequently, it is possible to fit the model to long-term epidemics experiencing multiple waves and peaks – some of the US cases, which have more erratic behavior, illustrate this property.

### 12.4. Predicting the future

The driving parameter of an SEIR model is the effective reproductive number. The model predictions depend exponentially on *R*(*t*). Thus we need to know the future value of *R*(*t*) to make reliable predictions. As such, it is maybe more useful to run scenarios with values of *R*(*t*) leading to either unstable *R*(*t*) > 1, neutral *R*(*t*) = 1, and stable evolution *R*(*t*) < 1 of the epidemics. These scenarios allow for predicting the severity of the pandemics under different regimes of interventions in different countries. We expect that societal and cultural diversity between countries also affect the *R*(*t*). Thus, some countries may need to implement stricter interventions than others, to keep *R*(*t*) below one.

Assuming a country has implemented sufficient measures to reduce *R*(*t*) below one, the system can realistically simulate the future decline of the epidemics under the current interventions. With time and more data, we also expect to use the system for quantifying the impact of various opening measures on *R*(*t*). We already know that the reopening of schools and kindergartens in Norway only led to a slight increase in *R*(*t*).

Many countries do not test all deaths for COVID-19. As a consequence, the number of reported deaths is highly uncertain and tends to underestimate the actual number of COVID-19 deaths. France lists only the COVID-19 patients who die in a hospital. Consequently, we multiply these numbers by an adjustment factor in the assimilation to correct for this bias. For some other countries, e.g., The Netherlands, Quebec, England, and The US, the estimated number of deaths is larger than the reported number. This overestimation also influences the forecasts. The various projections of the number of fatalities reflect this uncertainty. Thus, we need to interpret the observed COVID-19 deaths as rough estimates.

### 12.5. Short term predictability

The application for Quebec also assessed the quality of making short-term predictions (two weeks ahead), using the current estimates of *R*(*t*). We found a significant forecast skill, suggesting that it may be beneficial to use model predictions for evaluating the need and impact of implementing new interventions. Also, the short-term forecasts can be useful for planning the ICU capacity.

### 12.6. Which data to condition on?

In this work, we have mostly conditioned the SEIR model on the accumulated number of deaths and the current number of hospitalized. In some examples, we have also included derived or adjusted data of the accumulated amount of cases (e.g., for England and France).

An issue is that the basic SEIR model does not include hospitalized and deaths (see, e.g., the flow diagram in Figure 1). Thus, these observations are not straightforward to use, unless we assume to know (as we did here) the fraction of cases that die or go to the hospital. The registration of COVID-19 patients in the hospital may not be very accurate and using the number of ICU patients, as is done for The Netherlands, the time until hospitalization *τ*_*hosp*_, and the hospitalization rate *p*_s_ must be adjusted to reflect the difference between the two groups of patients.

Also, many patients in care homes for the elderly become severely ill and die without going to the hospital. When we assign uncertainties to the fraction of patients going to the hospital, we can obtain an excellent match to the observations of the number of deaths. If the case fatality rate *p*_f_ and the hospitalization rate *p*_s_ are unknown, it is not possible to constrain the SEIR model with the observed numbers of deaths and hospitalized. However, we can condition the SEIR model on the observed data by using prior estimates of these parameters.

The observed number of cases would be useful if they were unbiased observations since they relate directly to the variables in the SEIR model. We could, in theory, obtain reliable case data by testing the whole or an unbiased sample of the population. With such data, we would not only much better constrain the model. We could even accurately estimate the case fatality rate *p*_f_ and the hospitalization rate *p*_s_. When health institutes provide an official estimate of the number of cases, this is typically a rough estimate of the total infected population that is always larger than the number of positive tests and hence biased. Selective testing and too little testing inevitably creates a bias in these data. Nevertheless, our data-assimilation approach permits us to estimate the total number of infected as part of the assimilation output when we assimilate data on death and hospitalized. This posterior estimate of the number of infected, and its uncertainty, may then be used to evaluate the bias in the raw data.

We don’t know the total number of infected, so we don’t know the basic reproduction number. We do see the number of deaths and the number of hospitalized, but since we do not know the actual number of infected, the severely and fatally ill fractions are both unknown. Thus, to obtain unbiased data, we should test extensively and representatively across the entire population.

Still, in some of the examples we included the case data in the conditioning, but assuming that they only detect a specified fraction of the actual number of cases, we added a more substantial uncertainty. The effect is that we might obtain a more realistic figure of infections (e.g., in England). In some cases, e.g., for Norway and The Netherlands, we instead used the case data to constrain the initial conditions to provide predictions in agreement with the assumed number of infections.

## 13. Summary

We have demonstrated the use of an ensemble-based data assimilation system for modeling and predicting the development of the COVID-19 outbreak in 11 countries and states. We used an extended SEIR type model, and obtained realistic predictions of the epidemics, with uncertainty estimates, by conditioning the model ensemble on observed deaths and hospitalization numbers. The system clearly illustrates the inherent instability in infectious illnesses where the development of the disease primarily depends on the effective reproductive number, *R*(*t*). The data assimilation system provides for online and automated estimation and monitoring of *R*(*t*). Unfortunately, there is a lag of two-three weeks between infections and registered hospitalization and deaths. Thus, when one observes a too high value of *R*(*t*), it may already be too late to avoid another round of significant restrictions on society. It is, therefore, mandatory to reopen society slowly, step by step, and allow enough time to assess each step’s impact before initiating the next one. In this respect, the system will provide an online estimation of any changes to *R*(*t*), although with the delay related to the development time of the disease.

## Data Availability

All data used are public data regarding numbers of COVID related deaths and hospitalizations.
The data are available in the Github repository.

https://github.com/geirev/EnKF_seir

## Appendix A. ESMDA algorithm

As explained in [23, 24], ESMDA solves the standard ensemble smoother (ES) update equations using a predefined number of steps. In each recursive update, ESMDA inflates the measurement errors to reduce the impact of the measurements. With correctly chosen inflation factors, the ESMDA update precisely replicates the standard Ensemble Smoother (ES) in the linear case. When the model or observation operators are nonlinear, it turns out that the use of multiple short update steps reduces the errors and improves the solution as compared to using one long update step in ES.

It is easiest to derive ESMDA by using a tempering of the likelihood function in (16) [43], which leads to a recursive minimization of a sequence of *n* = 1, *N*_mda_ cost functions, [59, 23],

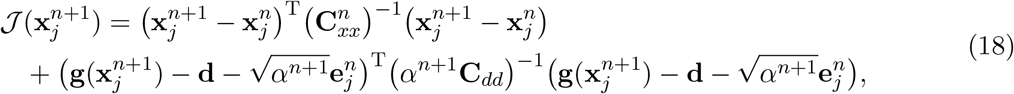

where we evaluate 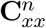 at the *n*th iterate **x**^*n*^, and we must have the following condition on the sequence of measurement-error inflation factors *α*^*n*^:

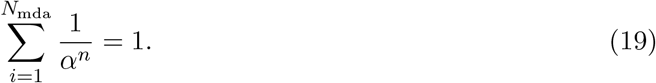

In each ESMDA step we must minimize *j* = 1, …, *N* cost functions, one for each of the *N* members of the ensemble.

Similarly to the derivation of the standard EnKF analysis update, we obtain the recursive update equations for ESMDA given by Eqs. (24) in the algorithm below. The update direction is computed based on a linearization around the prior realizations of each update step.

To compute the ESMDA solution, we start by sampling the initial ensembles from

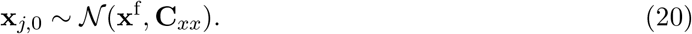

Then we integrate the model according to Eq. (14) to obtain the prior ensemble prediction for the first ESMDA step,

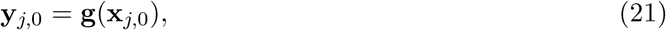

and we compute recursively the following for each iteration *n* = 0, …, *N*_mda_ − 1:

1. Construct the sample covariances 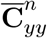 and 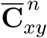, from the ensemble realizations 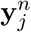 and 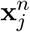.
2. Sample the measurement perturbations

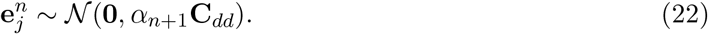
3. Generate the perturbed measurements

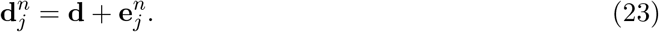
4. Compute the update

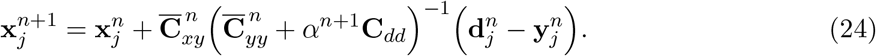
5. Rerun the model to obtain the updated prediction

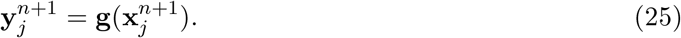

Repeat this procedure until *n* = *N*_mda_ −1, which results in the ESMDA solution for **x**_*j*_ and **y**_*j*_.

As in ES, the update direction is computed based on linearization around the current estimate of each update step. Thus, we can interpret the ES update as taking one long Euler step of length Δ*τ* = 1 in pseudo time *τ*, while in ESMDA we take a predefined number of shorter Euler steps of step length Δ*τ*_*i*_ = 1*/α*_*i*_ that satisfy Eq. (19) [23]. By updating the linearizations around each recursive estimate, ESMDA reduces the impact of nonlinearity and leads to improved solutions.

### A.1. Sensitivity of ESMDA to the number of steps

We have performed sensitivity experiments to examine the convergence properties of the ESMDA algorithm. We expect that the accuracy of the solution will improve with the number of steps, until a certain level where there is nothing more to gain. [23] examined the convergence of ESMDA for a simple nonlinear scalar case and obtained minimal improvement after 16-32 steps. Similar results were found here when examining the posterior solutions for deaths and hospitalized when using ESMDA with 1, 2, 4, 8, 16, 32, 64, and 128 steps, and similarly for the corresponding estimates of *R*(*t*). From visual inspection of the results, it is hard to justify more than 16 steps. We decided to use 32 steps in all the simulations presented in this manuscript to ensure convergence with a margin. A document with supplementary material including the results of the sensitivity tests is available from the Github repository https://github.com/geirev/EnKF_seir/doc/sensitivity.pdf.

### A.2. Sensitivity of ESMDA to the size of the ensemble

Finally, we have examined the convergence of ESMDA concerning the ensemble size. ESMDA, being based on a Monte Carlo algorithm, means that we can always improve the solution by increasing the ensemble size. However, we need to decide on a tradeoff between ensemble size, *N*, and the number of ESMDA steps, *N*_mda_, due to limitations on computing power. While the number of ESMDA steps impacts the actual convergence of the algorithm towards the correct solution, the ensemble size impacts the precision of the statistical estimate (i.e., how accurate we describe the error) of the final solution. We find it most important to first converge to the correct physical solution and, then, use an as large as possible ensemble to reduce the sampling errors. We examined the results using different ensemble sizes and two different random seeds. Even using *N* = 100 ensemble members, the posterior predictions are consistent with the data and very similar to the cases with larger ensemble sizes. There is a visual difference of the results on the random seed, which is more clear from the estimated *R*(*t*). When using 1000 or 5000 realizations, there is still a significant difference in the estimated *R*(*t*). Between 5000 and 10000 realizations, we could hardly see any difference in the parameter estimates or predictions. Thus, we use 5000 realizations in all the simulations in this paper. We have included the results in the supplementary material found at https://github.com/geirev/EnKF_seir/doc/sensitivity.pdf

### A.3. Observations

We have in most of the experiments conditioned the model on the measured deaths and hospitalization numbers. It is difficult to use the number of registered cases since these data are biased, as there is a capacity to test only a fraction of the total number of infected. However, we have used these data in some experiments assuming a fraction of positive tests over the real number of cases and using a significant measurement error. This approach sometimes helped to constrain the overall number of cases to “realistic” numbers.

### A.4. Uniqueness of the solution

Finally, we will comment on the degrees of freedom in the model parametrization versus the amount of information in the data. There are dependencies between several of the uncertain parameters. E.g., if we specify too few initial cases, this may be compensated for by a higher value of **R**(*t*) in the first period. If the model predicts too many infectious, then this can be compensated for by reducing the fractions *p*_s_ and *p*_f_. Thus the system appears to be under-determined. However, by specifying prior distributions for the uncertain parameters, we ensure one unique posterior solution, and we obtain it by running the model with the ensemble of estimated parameters as input.

## Acknowledgments

G. Evensen was supported by internal funding from NORCE (and his family was generous enough to allow and encourage him to work long days and nights given the urgency of this work). J. Amezcua, A. Carrassi, and A. Fowler were supported by the NERC award NCEO02004. F.C. Vossepoel was supported by a Delft Technology Fellowship. C. Jones and C. Sampson were supported by the US Office of Naval Research under grant N00014-18-1-2204. CEREA is a member of Institute Pierre-Simon Laplace (IPSL).

## Notes

### Competing Interest Statement

The authors have declared no competing interest.

### Funding Statement

All authors were funded either by their universities, research institutions, or they contributed with their own time.

### Author Declarations

No clinical data were used

